# Accuracy of diagnostic codes and algorithms used to identify rheumatoid arthritis and juvenile idiopathic arthritis in electronic health records: systematic review and meta-analysis

**DOI:** 10.1101/2025.06.30.25330552

**Authors:** Constanza Saka-Herrán, Jessica Bennett, Yara Alkabti, Muhammad Fatir, Barbara Clyne, Caroline McCarthy, Gráinne Tynan, Nikki Dunne, Michelle Flood, Eoghan McCarthy, Frank Moriarty

**Affiliations:** School of Pharmacy and Biomolecular Sciences, Royal College of Surgeons in Ireland (RCSI), Dublin, Ireland; School of Medicine, Royal College of Surgeons in Ireland (RCSI), Dublin, Ireland; Dept Public Health & Epidemiology, School of Population Health, Royal College of Surgeons in Ireland (RCSI), Dublin, Ireland; Department of General Practice, Royal College of Surgeons in Ireland (RCSI), Dublin, Ireland; Department of Rheumatology, Beaumont Hospital/Royal College of Surgeons in Ireland (RCSI), Dublin, Ireland

**Author notes:** **<u>Address</u>** School of Pharmacy and Biomolecular Sciences, Royal College of Surgeons in Ireland (RCSI) 1^st^ Floor Ardilaun House Block B, 111 St Stephen’s Green, Dublin 2, Ireland. **<u>Corresponding Author</u>** Frank Moriarty. **<u>Grant/Financial support</u>** This research was funded by the Health Research Board Applied Programme Grant Awards (APRO-2023-028) scheme.

## Abstract

**Objective:** This systematic review aimed to assess the diagnostic accuracy of algorithms used to identify rheumatoid arthritis (RA) and juvenile idiopathic arthritis (JIA) in electronic health records (EHRs).

**Methods:** We searched MEDLINE, Embase, and CENTRAL databases and included studies that validated case definitions against a reference standard such as rheumatologist-confirmed diagnosis or ACR/EULAR classification criteria. Title/abstract screening, full-text review, data extraction and quality assessment were all completed in duplicate. Results were synthesised narratively and using a bivariate random-effects meta-analysis of sensitivity and specificity.

**Results:** A total of 35 studies were included. Algorithms varied widely in complexity, ranging from single ICD codes to combinations including disease-modifying antirheumatic drugs (DMARDs), hospitalisation records, and specialist diagnosis. Algorithms combining ICD codes with DMARD prescriptions (pooled sensitivity= 0.79 95% CI 0.61-0.90, specificity= 0.96 95% CI 0.72-1.00, PPV= 0.78 95% CI 0.63-0.88) or requiring an ICD code assigned by a rheumatologist (pooled sensitivity= 0.91 95% CI 0.70-0.98, specificity= 0.94 95% CI 0.49-1.00, PPV= 0.70 95% CI 0.64-0.75) showed the highest accuracy, with balanced sensitivity, specificity, and positive predictive value (PPV). Less restrictive algorithms demonstrated high sensitivity but lower PPV. Substantial heterogeneity was observed across studies, likely due to differences in algorithm structure, data sources, and validation methods. Despite this variability, we used conceptually coherent categories to allow for meaningful synthesis, prioritising clinical interpretability.

**Conclusions:** These findings support the use of more specific algorithms when diagnostic certainty is essential and highlight the need for further validation of high-performing algorithms across diverse healthcare systems.

**Significance and Innovations:**

▪ This is the first comprehensive systematic review to evaluate and synthesize the accuracy of algorithms used to identify rheumatoid arthritis and juvenile idiopathic arthritis in electronic health records (EHRs), addressing a growing need as real-world data become increasingly central in rheumatology research.
▪ The findings provide critical guidance for researchers and clinicians on the strengths and limitations of commonly used case definitions, helping improve validity of studies using administrative or EHR data.
▪ By categorizing algorithms based on their components and reference standards, this review offers a practical framework for selecting the most appropriate algorithm depending on the study purpose and data source.
▪ The review highlight gaps in validation efforts and emphasizes the need to validate high-performing algorithms across diverse healthcare settings and evolving coding systems, ensuring accurate disease identification in current and future research.

## 1. Introduction

Rheumatoid arthritis (RA) and juvenile idiopathic arthritis (JIA) are autoimmune diseases characterised by joint inflammation, and systemic complications, leading to disability, and reduced quality of life^1^. Despite differences in onset and classification criteria, they share key pathogenic mechanisms, clinical manifestations and therapeutic strategies^2,3^. A challenge in RA/JIA research is obtaining robust epidemiological data to understand the natural history of disease and impact of therapy on disease outcomes^4^. Electronic health records (EHRs), have greatly facilitated studying these conditions in large populations, providing comprehensive data on patient demographics, symptoms, diagnoses, and treatment history^5,6^, in a real-world setting.

However, the reliability and validity of research based on EHR data depend on the accuracy of the diagnostic codes and algorithms used to identify the condition^7,8^. Combining multiple diagnostic codes with clinical data, such as prescriptions and laboratory results, enhances RA identification accuracy. For instance, algorithms incorporating at least three physician diagnostic codes, achieve specificity and positive predictive values (PPVs) of over 90%^9^. The inclusion of disease-modifying antirheumatic drug (DMARD) prescriptions with diagnostic codes, significantly improves case identification^10^. Standardised algorithms that integrate International Classification of Diseases (ICD) codes with other clinical parameters have also been assessed for their utility in diverse healthcare systems^11^.

Despite advances in identifying RA/JIA cases, limitations persist due to coding inaccuracy, population heterogeneity, and differences across healthcare systems and methodological approaches. A 2013 systematic review, part of the Mini-Sentinel pilot project sponsored by the Food and Drug Administration (FDA), reported substantial variability in the accuracy of algorithms (PPV 34%-97%) applied to EHRs^12^. The data sources of interest were limited to the US or Canada reflecting Mini-Sentinel’s focus on US-based data for surveillance system^13^, thus limiting the global applicability of the findings.

The aim of this systematic review is to synthesise and critically assess evidence on the accuracy of diagnostic codes and algorithms used to identify RA and JIA in EHRs and other administrative databases. By expanding the focus to include other regions, this review provides a more comprehensive evaluation of the tools used to identify the disease across various key data sources in the context of the significant changes in RA/JIA treatment and outcomes over the past decade.

## 2. Methods

The study protocol was registered on PROSPERO (Registration ID: CRD42024619130). The systematic review is reported in line with the PRISMA-DTA statement^14^.

### 2.1 Eligibility criteria

We included studies assessing the accuracy of codes and algorithms identifying RA and JIA in EHRs or other administrative databases against a “reference standard” based on clinician-documented diagnosis, clinician-conducted chart review, another form of clinician confirmed diagnosis, or application of the American College of Rheumatology/ European League Against Rheumatism (ACR/EULAR) criteria (e.g., to GP records, medical charts, office records) regardless of study design. Studies relying on self-reported diagnoses as the reference standard were excluded, as previously recommended^15^. Eligible studies had to clearly report algorithm components and describe methods for accuracy assessment. We excluded: case reports, editorials, commentaries, and narrative reviews. While systematic reviews were not included, relevant ones were used for reference checking. No restrictions were placed on language or publication date.

### 2.2 Information sources

We searched MEDLINE (PubMed interface), EMBASE and the Cochrane Central Register for Controlled Trials. Search strategies used medical subject headings (MeSH) terms and text-words aligned with our eligibility criteria (Supplementary Appendix 1) and were adapted for each database. To ensure literature saturation, we manually reviewed references of eligible studies and included articles from prior systematic reviews.

### 2.3 Search strategy

Our strategy was based on the Mini-Sentinel project search strategies^12,16^, and updated with additional terms. We reviewed the PubMed MeSH terms, as well as abstracts and full-texts of primary studies from a prior systematic review to identify relevant terminology. The search was developed collaboratively and validated by confirming it retrieved all articles from the Chung et al review^12^. After finalizing the MEDLINE search, it was adapted to the other databases. Full search strategies and validation process details are in Supplementary Appendix 2. Searches were conducted on November 11, 2024.

### 2.4 Study records

#### 2.4.1 Data management

We used Covidence software for record management and deduplication. Screening questions for title/abstract review were developed and tested. Reviewers completed training and pilot screening (50 abstracts by five reviewers) to calibrate eligibility criteria.

#### 2.4.2 Study selection

Two reviewers (of CSH, JB, YA, MuF, FM) independently screened titles/abstracts against eligibility criteria. Full-texts of potentially eligible studies were reviewed in duplicate. Disagreements were resolved by discussion or a third reviewer. Reasons for full-text exclusions were documented.

#### 2.4.3 Data collection process and items

Two reviewers independently extracted data using a standardized template based on the STARD criteria^17^, adapted for administrative databases. Discrepancies were resolved through discussion or a third reviewer. Extracted data included study characteristics, algorithms definitions, validation methods, and accuracy measures. The template is available as a data supplement (see availability of data statement). Classifications for target condition are provided in Supplementary Appendix 3.

### 2.5 Diagnostic accuracy measures

Primary accuracy measures were sensitivity, specificity, PPV, and negative predictive value (NPV). When not reported, these were calculated from two-by-two tables. For practical interpretation, accuracy was categorized is high (≥80%), moderate (60-80%), or low (<60%)^18^. The unit of assessment was the individual. Further details are provided in Supplementary Appendix 3.

### 2.6 Risk of bias and applicability

Quality was assessed using the QUADAS-2 tool, which examines risk of bias and applicability across four domains: patient selection, index test, reference standard, and flow/timing^19^. Domains were rated as low, unclear or high for each domain. Overall study bias followed QUADAS-2 guidance (i.e., any high-risk domain rendered the study high risk overall)^14^. Two reviewers independently assessed study quality, resolving disagreements through discussion.

### 2.7 Synthesis of results

We categorized algorithms based on the number and type of codes used to identify RA/JIA (e.g., ICD, prescriptions, diagnostic tests, procedures). Classification followed Shrestha et al^20^ with modifications:

▪ *Less restrictive algorithms*: required a single diagnostic code from an outpatient visit or unspecified source (e.g., single ICD code).
▪ *Restrictive algorithms*: required multiple codes or inclusion of specific types (e.g., procedures, prescriptions, tests, or hospitalisation codes).

Algorithms were categorised as either restrictive or less restrictive. For further analysis, we developed sub-categories among restrictive algorithms based on expected accuracy and usage frequency in included studies. These were: RA diagnosis codes and DMARD prescription codes, ≥2 ICD codes for RA, ≥1 ICD code for RA by rheumatologist, and ≥1 ICD hospitalisation code for RA (full description are in Supplementary Appendix 3).

### 2.8 Meta-analysis

We performed a bivariate random-effects meta-analysis using a generalised linear mixed-effects model (GLMM) to jointly synthesise sensitivity and specificity estimates, accounting for within- and between-study variability and their correlation^21–23^. Pooled PPV estimates were obtained via meta-analysis of proportions using the *meta* package in R, applying random-effects models within subgroups defined by algorithm and reference standard categories (*metaprop* function).

For each study, sensitivity and specificity with 95% CIs were calculated. Forest plots were generated for PPV, sensitivity, and specificity, displaying individual and pooled estimates by algorithm subgroup to capture between-study heterogeneity. Heterogeneity was assessed using I^2^ statistics derived from variance components of the random-effects model. Analyses were conducted using R (version 4.4.3).

## 3. Results

### 3.1 Study selection

The search yielded 4,109 records; after removing duplicates, 3,452 were screened based on eligibility criteria, and 108 underwent full-text review. Seventy-seven articles were excluded, reasons provided in Figure 1. Reference checks of the 31 eligible articles identified 4 additional studies, totalling 35 included in the review (Figure 1).

**Figure 1.**
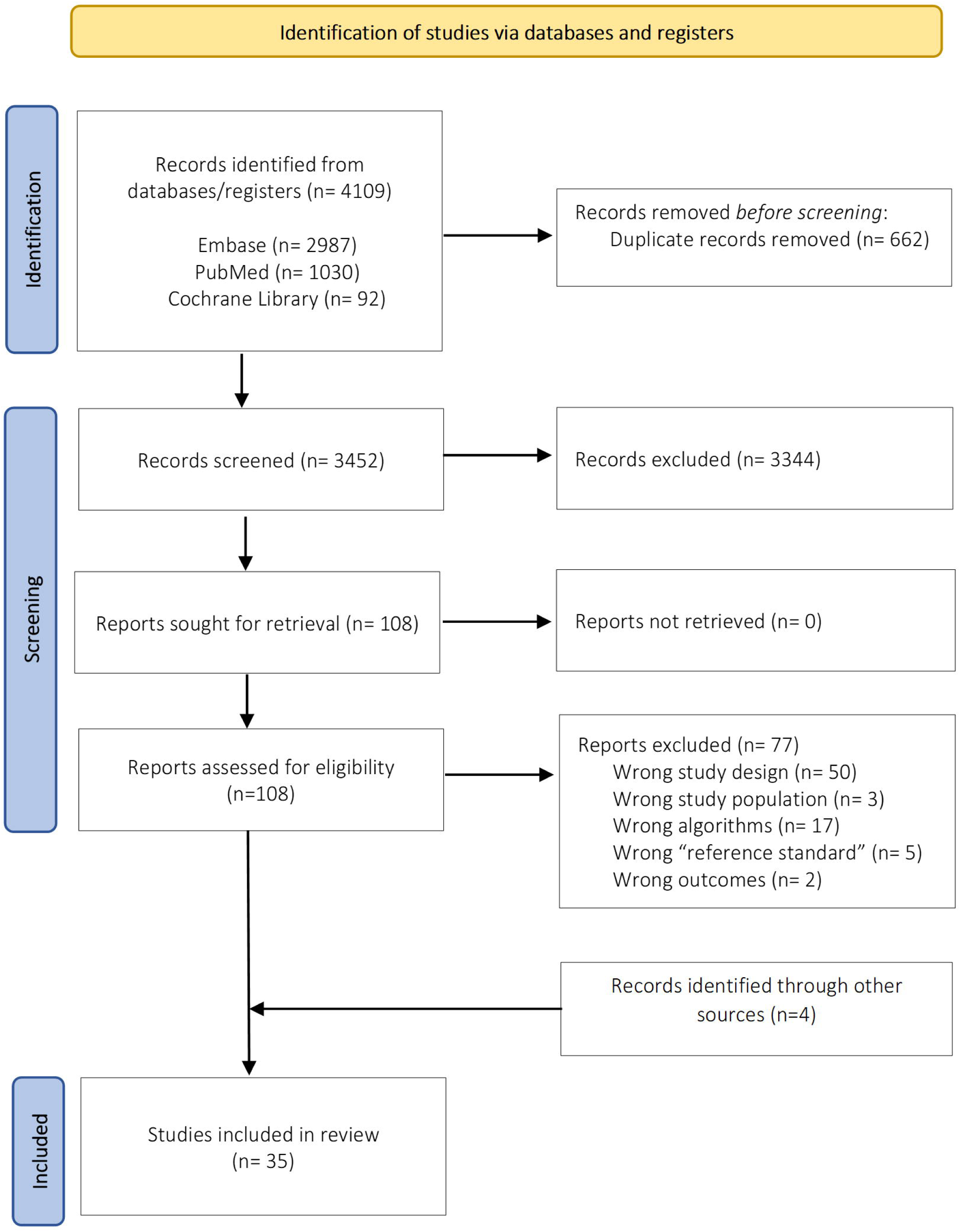
Flow diagram for study selection.

### 3.2 Study characteristics

Most studies were from the US and Canada (n=21), followed by Europe, (mainly Italy, UK, Denmark and Sweden; n=10), Asia (Japan and South Korea; n=3), and Australia (n=1). The majority were retrospective cohorts (n=25), followed by cross-sectional (n=6) and case-control studies (n=4). Two-thirds (n=23, 66%) were published since the 2013 review (Supplementary Appendix 4-Table 1).

**Table 1.** Summary characteristics and accuracy measures of included studies (n=35)

| Study | Codes/Algorithms |  |  |  |  | Reference standard |  |  | Measures of Accuracy / Results |  |  |  |
| --- | --- | --- | --- | --- | --- | --- | --- | --- | --- | --- | --- | --- |
|  | Restrictive |  |  | Less restrictive |  | CC | DR | MRR | Se | Sp | PPV | NPV |
|  | D | M | L | D | M |  |  |  |  |  |  |  |
| Rheumatoid Arthritis |  |  |  |  |  |  |  |  |  |  |  |  |
| Allebeck et al. 1983<sup>29</sup> |  |  |  | X |  | X | X |  | - | - | 56.5-65.2% | - |
| Almutairi et al. 2021<sup>51</sup> | X | X |  |  |  | X | X |  | 25-90% | 16.6-85.7% | 63.6-97.9% | 7.6-30% |
| Carrara et al. 2015<sup>11</sup> | X | X |  | X | X | X |  |  | 82.4-96.3% | 90.3-99.8% | 72.5-85% | 97.1-99.9% |
| Carroll et al. 2012<sup>32</sup> | X | X | X |  |  |  | X |  | 57-79% | - | 87-95% | - |
| Cho et al. 2013<sup>24</sup> | X | X |  |  |  | X |  |  | 84.7-96.5% | 30.8-70.0% | 87.3-94.0% | 32.9%-76.2% |
| Convertino et al. 2021<sup>53</sup> | X | X |  |  |  |  | X |  | 37-93% | 84-95% | 74-82% | 72-95% |
| Curtis et al. 2018<sup>25</sup> | X |  |  |  |  |  | X |  | - | - | 68-75% | - |
| Fowles et al. 1995<sup>26</sup> |  |  |  | X |  |  |  | X | 44.4% | 99.7% | 44.4% | 99.7% |
| Hanly et al. 2015<sup>44</sup> | X |  |  | X |  |  | X |  | 20.7-94.8% | 62.5-98.5% | 38.7-77.1% | 83.2-98.0% |
| Huang et al. 2020<sup>33</sup> | X | X | X |  |  | X | X |  | 47-89% | 50-95% | 54-93% | 50-90% |
| Ibfelt et al. 2017<sup>40</sup> | X |  |  |  |  |  | X |  | - | - | 79-92% | - |
| Katz et al. 1997<sup>41</sup> |  |  |  | X |  |  | X |  | 90% | - | 95% | - |
| Kim et al. 2011<sup>10</sup> | X | X |  |  |  | X | X |  | - | - | 33.6-88.9% | - |
| Kronzer et al. 2020<sup>38</sup> | X |  | X |  |  | X |  |  | 13-71% | 97-100% | 90-100% | 61-80% |
| Kubota et al. 2021<sup>52</sup> | X | X |  |  |  |  | X |  | 26-86.3% | 99.6-100% | 55.3-89.6% | 99.6-99.9% |
| Liao et al. 2010<sup>34</sup> | X | X | X |  |  | X | X |  | 51-63% | - | 88-94% | - |
| Linauskas et al. 2018<sup>30</sup> | X | X |  | X |  | X | X |  | - | - | 25-92.6% | - |
| Losina et al. 2003<sup>27</sup> | X |  |  |  |  |  |  | X | 65% | - | 86% | - |
| Nanji et al. 2012<sup>48</sup> |  |  |  | X |  |  | X |  | - | - | 70.9% | - |
| Ng et al. 2012<sup>49</sup> | X | X |  |  |  | X |  | X | 38.1-88.1% | 68.3-98.4% | 30.9-91.4% | - |
| Palta et al. 2021<sup>42</sup> | X | X | X |  |  |  | X |  | - | - | 71-98% | 100-100% |
| Pedersen et al. 2004<sup>31</sup> | X |  |  |  |  | X | X |  | - | - | 46-58.9% | - |
| Singh et al. 2004<sup>50</sup> | X | X | X | X |  |  | X |  | 76.5-100% | 55-97.1% | 66.2-97% | 77-86.2% |
| Sugiyama et al. 2024<sup>64</sup> | X | X |  |  |  | X | X |  | - | - | 22.6-88.6% | - |
| Thomas et al. 2008<sup>47</sup> | X | X |  |  |  | X |  |  | 78-93% | 40-96% | 64.8-96% | 69.5-84.5% |
| Waldenlind et al. 2014<sup>43</sup> | X |  |  | X |  | X |  |  | - | - | 83.3-91% | - |
| Wei et al. 2016<sup>36</sup> | X | X |  | X | X |  |  | X | 0-60% | - | 0-88% | - |
| Widdifield et al. 2013<sup>28</sup> | X | X |  | X |  |  | X |  | 23-100% | 60-96% | 55-93% | 72-100% |
| Widdifield et al. 2014<sup>9</sup> | X |  |  | X |  |  | X |  | 22-90% | 97-100% | 20-88% | 99-100% |
| Zheng et al. 2022<sup>37</sup> | X |  | X |  |  | X | X |  | 71.9-75.5% | 95.2-99.5% | 86.4-93.8% | 74.4-77.2% |
| Zhou et al. 2016<sup>65</sup> | X | X |  |  |  |  | X |  | 83-94% | 94.6-99% | 30.9-91.3 | - |
| Juvenile Idiopathic Arthritis |  |  |  |  |  |  |  |  |  |  |  |  |
| Harrold et al. 2013<sup>39</sup> | X |  | X |  |  |  | X |  | 81-100% | - | 45-91% | - |
| Peterson et al. 2023<sup>35</sup> | X |  |  |  |  |  | X |  | 63-100% | - | 48-97% | - |
| Stringer et al. 2015<sup>46</sup> | X |  |  | X |  | X | X |  | 53-86% | - | 52-65% | - |
| Thomas et al. 2008<sup>47</sup> | X | X |  |  |  | X |  |  | 40-100% | 0-100% | 86.2-100% | 21-80% |

Abbreviations: D= Diagnosis codes; M= Medication codes; L= Laboratory test codes; CC= Clinical Criteria (ACR, EULAR, ILAR); DR= Diagnosis or chart review by Rheumatologist; MRR= Medical Records Review by trained nurses or authors with no clear specification of the reference standard used; Se= Sensitivity; Sp= Specificity; PPV= Positive Predictive Value; NPV= Negative Predictive Value.

The included studies (n=35) used diverse electronic databases, including national health insurance claims-i.e. Korean National Health Insurance database^24^, Medicare^10,25–27^, and Ontario Health Insurance Plan^9,28^; hospital discharge records-i.e. Stockholm County Medical Information System^29^, Danish National Patient Registry^30,31^, Partners Healthcare database used by the Brigham and Women’s Hospital and the Massachusetts General Hospital^32–34^, and Canadian Institute for Health Information Discharge Abstract Database^9,28^; and institutional EHRs-i.e. Vanderbilt University Medical Center’s Synthetic Derivate^32,35,36^, University of California Los Angeles Health System^37^, Mayo Clinic Biobank^38^, and from specific rheumatological institutions or disease registries^11,39–41^. Some also incorporated pharmacy data ^30,42,43^, and physician billing records^41,44,45^.

Most studies focused on RA patients, though five examined JIA^35,39,45–47^. Samples sizes ranged from small cohorts, i.e., 31^48^ and 94^46^ participants, to large datasets exceeding 55,000 individuals^24^. Women comprised 50-80% of participants. Mean age varied, typically between 50-70 years for RA, and <19 years for JIA. Full study characteristics are detailed in Supplementary Appendix 4 - Table 1.

RA case identification relied mainly on ICD codes (Table 1), with ICD-9 (62.8%) and ICD-10 (48.6%) being the most used (Supplementary Appendix 4-Table 1). Some older datasets also used ICD-8 codes^29,30^. Additional criteria were applied in several studies, such as medications (54.3%), and laboratory markers (22.8%) like rheumatoid factor (RF) and anticyclic citrullinated peptide (anti-CCP) antibodies^34,49^ (Table 1). Some studies implemented complex algorithms, including the eMERGE algorithm, which combined ICD codes with RF values and other autoimmune disease exclusions^37,38^. The algorithm developed by Liao et al, incorporating ICD codes, prescriptions and laboratory values^34^ was validated in other settings by Carroll et al^32^, and Huang et al^33^ (Supplementary Appendix 4-Table 1).

Most studies (71.4%) used a rheumatologist’s clinical diagnosis as the reference standard (Table 1), while others applied established classification criteria including the 1987 ACR criteria, 2010 ACR/EULAR criteria, and for JIA, the 2001 International League of Associations for Rheumatology (ILAR) criteria (Supplementary Appendix 4-Table 1). Four studies relied on medical records review by trained nurses or researchers, without clear specification of the reference standard^26,27,36,49^ (Table 1).

### 3.3 Risk of bias and applicability

Risk of bias and applicability concerns varied across studies, with patient selection and flow/timing being the most affected domains (Table 2). Patient selection showed the highest risk, mainly due to non-random inclusion, use of ICD codes without additional verification, and concerns regarding representativeness. The index test domain showed a low risk, generally due to clearly defined and consistently applied algorithms, though some studies lacked clarity or did not report blinding to the reference standard. The reference standard domain showed the lowest risk, using established standards like rheumatologist diagnosis or ACR/EULAR criteria, though some had unclear validation methods. Flow and timing had mixed results, with high risk in studies that did not include all participants in analysis or did not assess all diagnostic accuracy measures due to study design.

**Table 2.** Risk of bias and applicability concerns of included studies (QUADAS-2 tool).

| Author/Year/Country | Domain 1<br>Patient Selection |  | Domain 2<br>Index Test |  | Domain 3<br>Reference standard |  | Domain 4<br>Flow and Timing |
| --- | --- | --- | --- | --- | --- | --- | --- |
|  | Risk | Concern | Risk | Concern | Risk | Concern | Risk |
| Allebeck et al, 1983, Sweden | High | Unclear | Unclear | Unclear | High | High | High |
| Almutairi et al, 2021, Australia | High | Unclear | Low | Low | Low | Low | High |
| Carrara et al, 2015, Italy | Unclear | Low | Low | Low | Low | Low | Unclear |
| Carroll et al, 2012, USA | Low | Low | Low | Low | Unclear | Low | Unclear |
| Cho et al, 2013, South Korea | Unclear | Unclear | Low | Low | Low | Low | Low |
| Convertino et al, 2021, Italy | Unclear | Low | Low | Low | Low | Low | Low |
| Curtis et al, 2018, USA | High | High | High | High | Low | Low | High |
| Fowles et al, 1995, USA | High | High | High | Unclear | High | High | Unclear |
| Hanly et al, 2015, Canada | Unclear | Unclear | Low | Low | Low | Low | Low |
| Harrold et al, 2013, USA | Unclear | Low | Unclear | Unclear | Unclear | Unclear | Unclear |
| Huang et al, 2020, USA | Unclear | Low | Low | Low | Low | Low | Unclear |
| Ibfelt et al, 2017, Denmark | High | High | Unclear | Unclear | Low | Low | High |
| Katz et al, 1997, USA | High | High | Unclear | Unclear | Low | Low | Unclear |
| Kim et al, 2011, USA | High | High | Low | Low | Low | Low | Unclear |
| Kronzer et al, 2020, USA | Unclear | Unclear | Low | Low | Unclear | Unclear | Low |
| Kubota et al, 2021, Japan | Unclear | Low | Low | Low | Low | Low | Unclear |
| Liao et al, 2010, USA | Low | Low | Low | Low | Low | Low | Unclear |
| Linauskas et al, 2018, Denmark | Unclear | Low | Low | Low | Unclear | Unclear | Unclear |
| Losina et al, 2003, USA | High | High | Unclear | Low | Unclear | Low | High |
| Nanji et al, 2012, Canada | High | High | Unclear | Unclear | Low | Low | High |
| Ng et al, 2012, USA | Unclear | Unclear | Low | Low | Unclear | Low | Unclear |
| Palta et al, 2021, Finland | Unclear | Low | Unclear | Unclear | Unclear | Unclear | Unclear |
| Pedersen et al, 2004, Denmark | Unclear | Low | Low | Low | Low | Low | High |
| Peterson et al, 2023, USA | Unclear | Low | Low | Low | Unclear | Unclear | Unclear |
| Singh et al, 2004, USA | Low | Low | Low | Low | Low | Low | Unclear |
| Shiff et al, 2017, Canada | Unclear | Low | Low | Low | Low | Low | Unclear |
| Stringer et al, 2015, Canada | Unclear | Low | Unclear | Unclear | Unclear | Unclear | Unclear |
| Sugiyama et al, 2024, Japan | Unclear | Unclear | Low | Low | Low | Low | Unclear |
| Thomas et al, 2008, UK | Low | Low | Unclear | Unclear | Low | Low | High |
| Waldenlind et al, 2014, Sweden | Unclear | Unclear | Unclear | Unclear | Low | Low | High |
| Wei et al, 2016, USA | Unclear | Unclear | Low | Low | Unclear | Unclear | Unclear |
| Widdifield et al, 2013, Canada | Unclear | Unclear | Low | Low | Low | Low | Low |
| Widdifield et al, 2014, Canada | Low | Low | Low | Low | Unclear | Unclear | Low |
| Zheng et al, 2022, USA | Low | Low | Low | Low | Unclear | Unclear | Unclear |
| Zhou et al, 2016, UK | Low | Low | Unclear | Unclear | Unclear | Unclear | Unclear |

Overall, six studies were rated high risk of bias^25–27,29,40,48^ due to limitations in multiple domains, potentially leading to misclassification and reduced generalisability. In contrast, studies by Carroll et al^32^, Cho et al^24^, Liao et al^34^, Singh et al^50^, and Widdifield et al 2014^9^ had low risk, with strong design across all domains (Table 2.)

Applicability concerns were generally low, suggesting good relevance to real-world settings. However, studies using specialised databases or narrow inclusion criteria (i.e., paediatric populations, single-centre databases, incident RA cases) had moderate concerns as they may not generalise to broader RA populations (Table 2).

### 3.4 Results of individual studies

#### 3.4.1 Rheumatoid Arthritis codes and algorithms

Individual study results are detailed in Supplementary Appendix 5-Table 2. Across studies, over 180 definitions/algorithms were evaluated. Percentage of studies that used less restrictive, restrictive algorithms and their categories are presented in Figure 2.

**Figure 2.**
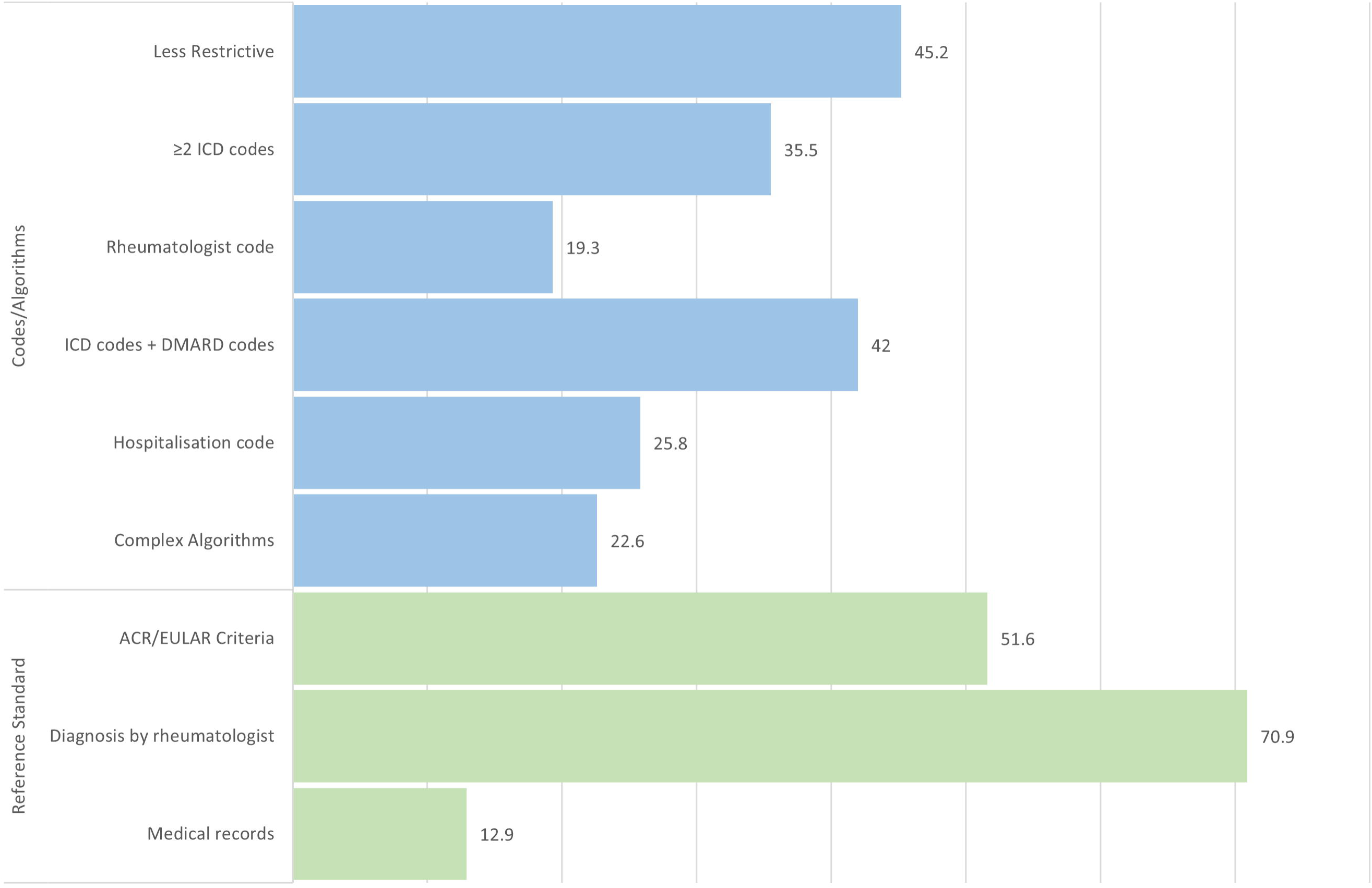
Codes/algorithms and reference standards used by the included studies assessing rheumatoid arthritis in electronic health records, % (n=31).

Less restrictive algorithms typically had high sensitivity but low specificity, increasing false positives. For example, Almutairi et al^51^ found 90% sensitivity but only 28.5% specificity using ≥1 RA primary code in hospital discharge records, highlighting the risk of over-identification. Other studies^28,44,50^ showed similar trends, with sensitivities >90% and specificities below 65%.

Adding DMARD prescriptions to ICD codes improved specificity and PPV while maintaining good sensitivity. Cho et al^24^ achieved 85% sensitivity and 70% specificity using ICD-10 codes with biologic, DMARD, or NSAID prescriptions, maintaining >85% PPV values in all developed algorithms. Similarly, Singh et al^50^ reported 85% sensitivity, 83% specificity, and 81% PPV when using a similar approach. Linauskas et al^30^ showed PPV increased from 61.9% to 87.7% when requiring ≥1 DMARD prescription.

Requiring ≥2 ICD RA codes, separated over time, improved specificity while maintaining good sensitivity, though decreasing PPV. Hanly et al^44^, reported 83% sensitivity, 81% specificity, and 52% PPV using two RA physician codes ≥2 months apart. Similar trends were seen in Kubota et al^52^ (84% sensitivity, 99% specificity, and 59% PPV) and Widdifield et al^9^ (84% sensitivity, 99% specificity, and 46% PPV).

Requiring ≥1 hospitalisation record improved specificity and PPV but greatly reduced sensitivity. Convertino et al^53^ found 95% specificity, 81% PPV but only 37% sensitivity when requiring 1 RA hospitalisation or emergency admission code. Similarly, Hanly et al^44^ achieved 98.5% specificity, 77% PPV and 20.7% sensitivity when using ≥1 hospitalisation code. Similar patterns were noted by Wididifield et al^9,28^.

Algorithms requiring at least one rheumatologist diagnosis code showed overall moderate to high sensitivity and specificity, with moderate PPV. Widdiefield et al 2014^9^ and 2013^28^, achieved 81% and 99% sensitivity, 99% and 77% specificity and 51% and 68% PPV, respectively. Similarly, Hanly et al^44^ reported 88% sensitivity, 75.5% specificity, and 47% PPV when using ≥1 RA code by a rheumatologist. In contrast, Carrara et al^11^ reported 99.8% specificity but only 13% sensitivity when combining one RA rheumatologist certification with other ICD-9 codes.

The eMERGE algorithm-combining ICD codes, RF values, and other autoimmune disease exclusions-demonstrated high specificity (95-99%) and PPV (94-97%) in both studies evaluating it (Zheng et al 2022^37^ and Kronzer et al 2020^38^, respectively). However, sensitivity varied likely due to differences in sample size and study population. Zheng et al^37^ (n=4,766, UCLA Health System) reported a higher sensitivity (72%), while Kronzer et al^38^ (n= 497, Mayo Clinic Biobank included in the Rochester Epidemiology Project) achieved a sensitivity of 53% in a smaller and more geographically limited sample. However, Kronzer et al^38^ found sensitivity increased with RA duration and EHR history, reaching 71% for patients with ≥10 years of disease. Overall, the eMERGE algorithm performed well, with sensitivity maybe influenced by dataset scope.

The RA algorithm initially developed by Liao et al^34^, validated by Carroll et al^32^ and Huang et al^33^, consistently showed moderate sensitivity (51-79%) and high PPV (>87%) across different settings showing a good balance between measures for identifying RA cases. Huang et al^33^ enhanced the model by incorporating ICD-10 codes and newer biologic treatments, achieving 77% sensitivity, 95% specificity, and 91% PPV. Overall, the algorithm maintains high accuracy, with sensitivity improving slightly as case definitions and coding systems evolved.

Both the eMERGE and Liao algorithms used laboratory data (RF, anti-CCP), which improved specificity and PPV, and had limited effect on sensitivity. Paltta et al^42^ reported 98% PPV when incorporating anti-citrullinated protein antibodies (ACPA) positivity in their algorithm, compared with ICD codes alone (82%) or ICD codes and DMARDs (89%). Singh et al also showed improved sensitivity (88%), specificity (91%), and PPV (93%) when adding positive RF to their algorithm, compared to ICD codes alone (sensitivity: 100%; specificity: 55%; PPV: 66%) or ICD codes and DMARDs (sensitivity: 85%; specificity: 83%; PPV: 81%)^50^, suggesting that laboratory data helps reduce false positives while maintaining a balanced improvement in the accuracy measures.

#### 3.4.2 Juvenile Idiopathic Arthritis codes and algorithms

JIA studies evaluated similar EHR-based algorithms (Supplementary Appendix 5-Table 2). Less restrictive algorithms, like ≥1 JIA diagnosis code, showed high sensitivity but moderate to low PPV. Harrold et al^39^ and Peterson et al^35^ both reported 100% sensitivity with PPVs of 45% and 48%, respectively.

Only one study assessed the accuracy of DMARD prescription alone as a definition. Thomas et al^47^ found 40% sensitivity, but 100% specificity and PPV when requiring ≥1 DMARD prescription without prior alternative indication.

Requiring ≥2 ICD codes for JIA improved PPV while maintaining good sensitivity. Reported sensitivity ranged from 72-93% and PPV from 60-100% across studies^35,39,45–47^. Peterson et al^35^ noted that increasing the number of JIA codes (≥4) raised PPV (83%) without substantial decrease in sensitivity (87%), compared to using ≥1 code (PPV=48%). Shiff et al^45^ found specificity improved (85-92%) with more ICD codes over longer intervals.

Two studies assessed algorithms requiring ≥1 JIA code from a rheumatologist. Harrold et al^39^ showed that requiring a rheumatologist visit improved PPV from 69% to 90% while maintaining good sensitivity (81%). Stringer et al^46^ showed similar sensitivity (86%) but decreased PPV (58%), which increased to 65% when using two JIA codes ≥8 weeks apart within 2 years, maintaining good sensitivity (81%).

### 3.5 Meta-analysis

Meta-analyses showed substantial variability in the accuracy of RA identification algorithms in EHRs, depending on algorithms components and reference standards. Using rheumatologist diagnosis as reference, algorithms requiring ≥2 ICD codes had the highest pooled sensitivity (0.89 95% CI 0.75-0.95, n=4 studies) and specificity (0.96 95% CI 0.74-1.00, n=4), though with high heterogeneity (I^2^= 91.3%, I^2^= 99.8%, respectively). Algorithms combining ICD codes with DMARD prescriptions also showed good pooled sensitivity (0.79 95% CI 0.61-0.90, I^2^= 90.6%, n=5) and high specificity (0.96 95% CI 0.72-1.00, I^2^= 98.9%, n=5) (Table 3, Supplementary Appendix 6-Figure 1). Algorithms using ≥1 ICD code by rheumatologist also showed high performance (sensitivity 0.91, 95% CI 0.70-0.98; specificity 0.94, 95% CI 0.49-1.00, n=3), but were based on only three studies, warranting cautious interpretation. A meta-analysis of sensitivity and specificity using ACR/EULAR classification criteria as the reference standard was not feasible due to insufficient data.

**Table 3.**
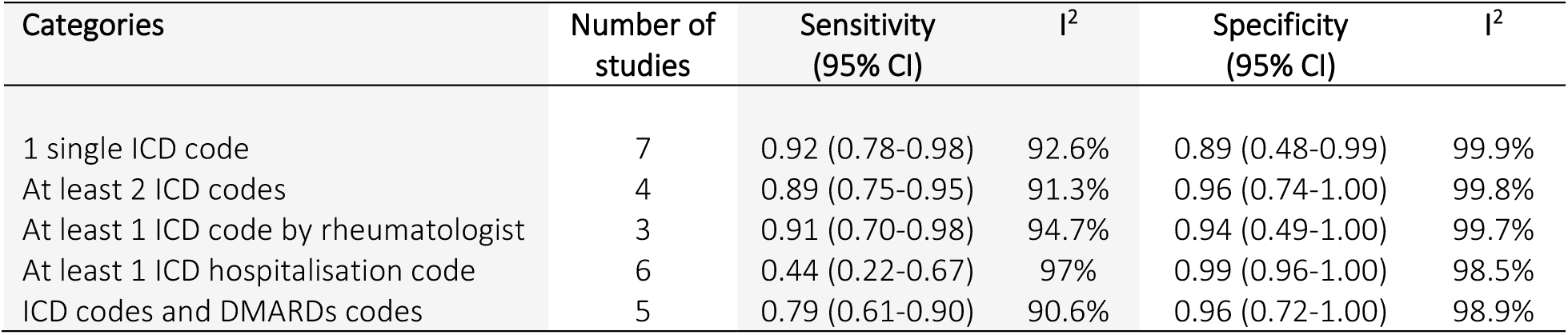
Pooled estimates of sensitivity and specificity and across algorithms categories (reference standard= diagnosis by rheumatologist).

PPV results varied by reference standard and showed substantial heterogeneity. Using rheumatologist diagnosis as the reference, the highest pooled PPVs were seen in algorithms combining ICD and DMARDs codes (0.78 95% CI 0.63-0.88, n=10), those including ≥1 ICD code by rheumatologist (0.64 95% CI 0.51-075, n=4), and those requiring ≥1 hospitalisation code (0.79 95% CI 0.77-0.81, n=7). In contrast, when using ACR/EULAR classification criteria, PPVs were slightly lower when requiring ≥2 ICD codes (0.62 95% CI 0.14-0.94, n=2) or ICD and DMARDs codes (0.73 95% CI 0.45-0.90, n=6). Algorithms requiring ≥1 ICD code by rheumatologist demonstrated the highest pooled PPV (0.85 95% CI 0.35-0.98, n=3), though with high heterogeneity (I^2^= 96.3%) (Table 4, Supplementary Appendix 7– Figure 2).

**Table 4.** Pooled estimates of PPV across algorithms categories by reference standards.

| Categories | Diagnosis by rheumatologist |  |  | ACR/EULAR criteria |  |  |
| --- | --- | --- | --- | --- | --- | --- |
|  | Number of studies | PPV (95% CI) | I<sup>2</sup> | Number of studies | PPV (95% CI) | I<sup>2</sup> |
| 1 single ICD code | 14 | 0.63 (0.49-0.75) | 97.8% | 6 | 0.74 (0.54-0.87) | 92.6% |
| At least 2 ICD codes | 7 | 0.64 (0.52-0.75) | 94.5% | 2 | 0.62 (0.14-0.94) | 98.2% |
| At least 1 ICD code by rheumatologist | 4 | 0.64 (0.51-0.75) | 94.8% | 3 | 0.85 (0.35-0.98) | 96.3% |
| At least 1 ICD hospitalisation code | 7 | 0.79 (0.77-0.81) | 8.9% | - | - | - |
| ICD codes and DMARDs codes | 10 | 0.78 (0.63-0.88) | 96.5% | 6 | 0.73 (0.45-0.90) | 99.6% |

## 4. Discussion

### 4.1 Summary of evidence

This systematic review analysed the accuracy of algorithms for identifying RA and JIA in EHRs. Overall, algorithms combining ICD codes with DMARD prescriptions, or those that include ≥1 ICD code assigned by rheumatologist, demonstrated the highest accuracy, balancing sensitivity, specificity, and PPV. These algorithms are more reliable for identifying confirmed RA cases, though rheumatologist-assigned codes may miss cases diagnosed in primary care, potentially leading to underestimation of disease prevalence. Less restrictive algorithms are useful for initial case identification (highly sensitive) but may require additional criteria to enhance specificity and PPV. Requiring ≥2 ICD codes is a well-balanced approach, reducing false positives (higher specificity) without significantly lowering sensitivity, though achieving moderate PPV. Hospitalisation-based codes are highly specific for identifying severe RA cases but may exclude many non-hospitalised, mild/moderate cases, making it less suitable for general RA case identification.

For JIA, a single ICD code is highly sensitive but may overestimate prevalence due to false positives. Requiring ≥2 spaced ICD codes improves accuracy and seems to be the best approach, providing a good balance of sensitivity, specificity and PPV in EHRs or administrative data. However, validation evidence for JIA remains limited, highlighting the need for further research.

Our findings align with and extend previous systematic reviews on RA and connective tissue diseases algorithms in EHRs. Compared to the 35 studies we included, the 2013 review by Chung et al^12^ included 9 studies and found that algorithms combining diagnostic and prescription data-particularly those including specialist codes, medication or laboratory data-achieved higher PPVs (>80%), while ICD-only algorithms had more variable performance across settings, generally with low PPVs. Similar trends were seen in studies for other conditions, but generally lower PPVs (<65%) in general populations^20,54^, probably reflecting the challenges of identifying these conditions due to non-specific coding and overlap with other autoimmune conditions. Similarly, a systematic review by Crossfield et al^55^ on gout also highlighted variability in methods and performance, reflecting a lack of standardisation in defining rheumatologic conditions using real-world data. Compared to these conditions, RA appears to be more accurately identified in EHRs, especially when algorithms include medication data or are supported by specialist confirmation.

The accuracy of hospitalisation-based algorithms must be interpreted within the context of evolving healthcare delivery. RA hospitalisations have declined significantly over the past two decades due to earlier diagnosis, improved outpatient care, and widespread use of biologic therapies^56–58^. As a result, performance of algorithms using hospitalisation codes in historic data may be less representative of current patient populations. Similarly, treatment patterns have evolved, with earlier DMARD initiation, combined therapies (e.g., biologics and DMARDs)^59–61^, and the development of targeted-synthetic DMARDs (e.g., JAK inhibitors), which may affect the predictive value of medication-based algorithms applied to contemporary prescribing data.

The transition from ICD-9 to ICD-10 introduced more specific diagnosis codes, potentially influencing algorithm performance. In this review, 62.8% of studies examined ICD-9 and 48.6% examined ICD-10 codes, reflecting variability in study periods, country-level implementation, differences in capacity to adopt new coding systems, and the use of historical data sources. Algorithms developed with ICD-9 codes may not translate directly to ICD-10 without re-validation^62,63^, particularly for specific or rare disease subtypes. These changes underscore the need for regular update and validation of algorithms to align with current coding systems and clinical practice.

From a practical perspective, the selection of an algorithm for identifying RA or JIA should be guided by the purpose of the study and the nature of the data source. For studies aiming to evaluate treatment outcomes, assess adverse events, or conduct comparative effectiveness studies, algorithms with high specificity may be preferable to ensure the correct identification of true cases. In contrast, for surveillance or studies assessing prevalence or burden of disease, algorithms prioritising higher sensitivity may be preferable, even at lower PPV.

Future research should focus on validating existing algorithms, such as the eMERGE^37,38^ and Liao et al^34^ algorithms, across diverse healthcare systems and populations. While both show consistent diagnostic accuracy in diverse settings, the generalisability of the Liao algorithm beyond academic health centres, remains uncertain. Validation using community-based data is needed to ensure broader applicability. Clear and transparent reporting of algorithm components is essential for replication, external validation, and broader implementation in different contexts. Additionally, the development and validation of algorithms using machine learning or natural language processing techniques hold promise for further improving case ascertainment, particularly in settings where structured coding is limited.

### 4.2 Limitations

The studies reviewed exhibited substantial methodological variability, making direct comparisons difficult. Differences in sample size, sampling methods, settings, population, and data sources likely contributed to the high statistical heterogeneity in several analyses. Algorithm definitions varied widely, even within similar categories, highlighting a lack of standardisation in algorithm development and validation methods. Despite this, meta-analyses were feasible by grouping algorithms into broad, but conceptually coherent categories, prioritising simplicity and clinical interpretability. While many incorporated medication use, most studies grouped DMARDs as a single category, precluding an evaluation of the contribution of specific drugs or therapeutic classes to the accuracy. Several studies did not report full accuracy measures due to sampling designs, which involved evaluating only algorithm-positive cases, and thus only the PPV. The PPV depends on disease prevalence within the population being studied. Several studies selected their populations from hospital settings or rheumatologic clinics, where the prevalence of RA is likely to be higher than in the general population, potentially leading to an overestimation of the PPVs. Another source of variability may be the data used in each study and purpose for which it was originally generated, i.e., for claims/reimbursement versus clinical care. Therefore, findings should be interpreted cautiously, and ideally, validated in general populations to better understand their performance across varying levels of disease prevalence. Finally, the search focused on studies explicitly reporting diagnostic validation, potentially excluding relevant studies that addressed this in broader investigations (i.e., assessed an algorithm accuracy as an intermediate objective as part of an EHR-based study of RA outcomes). A protocol deviation was the omission of grey literature search due the nature of the review and to the expectation that relevant studies would be published primarily in peer-reviewed journals.

This systematic review incorporates comprehensive and contemporary evidence from diverse geographic areas. To our knowledge, is the first systematic review to comprehensively synthesise and quantitatively analyse RA and JIA algorithm accuracy in EHRs without geographic restriction across diverse healthcare settings. It adheres to PRISMA guidelines, systematically categorised algorithms by restrictiveness to facilitate clinically meaningful comparisons, and used the QUADAS-2 tool for transparent quality assessment.

### 4.3 Conclusions

This review highlights the variability in accuracy of algorithms used to identify RA and JIA in EHRs. Algorithms combining diagnostic codes and prescription data, particularly DMARDs, or a rheumatologist diagnosis demonstrated the highest overall performance, with a good balance of sensitivity, specificity, and PPV. Less restrictive algorithms may aid initial case identification but require refinement for specific research purposes. The choice of algorithm should be tailored to the study objective and data source. Future efforts should prioritise external validation of high-performing algorithms across diverse settings to improve standardisation and reliability in real-world data research.

## Data Availability

This research is based on published literature. All data used in this research are already included in the article or the supplementary material. The data extraction template is available at the Open Science Framework: [OSF | Data Extraction Template.xlsx](https://osf.io/xfpzg/).

## Acknowledgment

The authors acknowledge the use of large language model to assist with English grammar and spelling during the preparation of the manuscript.

## Availability of data

This research is based on published literature. All data used in this research are already included in the article or the supplementary material. The data extraction template is available at the Open Science Framework: <u>OSF | Data Extraction Template.xlsx</u>.

## Notes

**<u>Declaration of interests</u>** Authors declare no potential conflict of interests.

### Competing Interest Statement

The authors have declared no competing interest.

### Clinical Protocols

https://www.crd.york.ac.uk/PROSPERO/view/CRD420251056943

