## Supplementary Material for "Accuracy of diagnostic codes and algorithms used to identify rheumatoid arthritis and juvenile idiopathic arthritis in electronic health records: systematic review and meta-analysis"

### Supplementary Appendix 1. Search Terms

| | | |
|---|---|---|
| Population | Rheumatoid Arthritis terms | “Arthritis, Rheumatoid” [Mesh] OR<br>“Arthritis, Juvenile” [Mesh] OR<br>“Felty Syndrome” [Mesh] OR<br>“Caplan Syndrome” [Mesh] OR<br>(("Arthritis, Rheumatoid"[Mesh]) AND "Lung Diseases, Interstitial"[Mesh]) OR<br>“Rheumatoid Arthritis” [Text word] OR<br>“Rheumatoid Vasculitis” [Text word] OR<br>“Rheumatoid Nodule” [Text word] OR<br>“Rheumatoid Nodules” [Text word] OR<br>“Juvenile Idiopathic Arthritis” [Title/Abstract: ~3] OR<br>“Juvenile Chronic Arthritis” [Title/Abstract: ~3] OR<br>“Juvenile Rheumatoid Arthritis” [Title/Abstract: ~3] OR<br>“Felty Syndrome” [Text word] OR<br>“Caplan Syndrome” [Text word] OR<br>(("Rheumatoid Arthritis") AND ("Interstitial Lung Disease")) OR<br>“RA-ILD” [Text word] OR<br>“(rheumat* AND arthritis)” |
| Data source | Database terms | “Database” [Publication type] AND<br>“Databases, Factual” [Mesh] OR<br>“Database Management Systems” [Mesh] OR<br>“Medical Records Systems, Computerized” [Mesh] OR<br>“Electronic Health Records” [Mesh] OR<br>“Medical Record Linkage” [Mesh] OR<br>“Administrative Claims, Healthcare” [Mesh] OR<br>“Hospital Records” [Mesh] OR<br>“Patient Discharge” [Mesh] OR<br>“Electronic Data Processing” [Mesh] OR<br>“Automatic Data Processing” [Text word] [Title/Abstract: ~3] OR<br>“Insurance database” [Text word] [Title/Abstract: ~2] OR<br>“Record linkage” [Text word] [Title/Abstract: ~2] OR<br>“Data Warehouse” [Text word] [Title/Abstract: ~2] OR<br>“Registered Person Database” [Text word] [Title/Abstract: ~3] OR<br>“Electronic Medical Records” [Text word] [Title/Abstract: ~3] OR<br>(“Billing” AND (“Data” OR “Claims” OR “Codes”)) [Title/Abstract: ~3] OR<br>(“Claims” AND (“Administrative” OR “Data*”)) [Title/Abstract: ~3] OR<br>((“Administrative” OR “Healthcare”) AND (“Data*” OR “Records”) OR<br>“Administrative Data*” [Text word] OR<br>“Administration Database” [Text word] OR<br>“Physician claims” [Text word] OR<br>“Hospital Billing” [Text word] |

| | | |
|---|---|---|
| Index classification | Definition/Algorithm terms | “Algorithm*” [Mesh] OR<br>“Current Procedural Terminology” [Mesh] OR<br>“International Classification of Diseases” [Mesh] OR<br>“Diagnostic Algorithm*” [Text word] OR<br>“Classification Algorithm*” [Text word] OR<br>“Claim Based Algorithm*” [Text word] OR<br>“Diagnostic codes” [Text word] OR<br>“Diagnostic Definition*” [Text word] OR<br>“ICD codes” [Text word] OR<br>“ICD” [Text word] OR<br>“ACR Criteria” [Text word] |
| Outcome measures | Validation/Accuracy terms | “Validation Study” [Publication type] AND<br>“Sensitivity and Specificity” [Mesh] OR<br>“Predictive Value of Tests” [Mesh] OR<br>“Reproducibility of Results” [Mesh] OR<br>“Data Accuracy” [Mesh] OR<br>“ROC Curve” [Mesh] OR<br>“Receiver Operating Characteristic Curve” [Title/Abstract: ~4]<br>“Predictive value” [Text word] OR<br>“Diagnostic Accuracy” [Title/Abstract: ~3] OR<br>“Positive Predictive Value” [Title/Abstract: ~3] OR<br>“Negative Predictive Value” [Title/Abstract: ~3] OR<br>“Accuracy” [Text word] OR<br>“Sensitivity” [Text word] OR<br>“Specificity” [Text word] OR<br>“Validity” [Text word] OR<br>“Validation Process” [Text word] OR<br>“Case Ascertainment” [Text word] OR<br>“Diagnostic performance” [Text word] |

### Supplementary Appendix 2. Search strategy and validation process

In an attempt to validate our search strategy, we conducted a first search in MEDLINE to determine if we could retrieve all the articles included in the previous systematic review, combining the terms related with administrative databases, definitions/algorithms and validation/accuracy measures with an AND. As we did not identify all relevant studies, we combined the administrative databases and definitions/algorithms terms with an OR to build a more sensitive search. With this search strategy we were able to retrieve all relevant articles, with the exception of those studies that did not answer our PICO question directly, i.e. whose main objective was different from validating an algorithm for identifying rheumatoid arthritis in EHRs (e.g. studies that evaluate risk or incidence rates as their main objective). We reviewed these studies (those that could be accessed in full-text) to identify terms that would allow us to retrieve them in a new search. The terms identified included 'confirm*', 'verified' 'verification' 'identified', 'valid*' and 'definite diagnosis' which were incorporated to the validation/accuracy measures search strategy. We repeated the first process retrieving all relevant studies but without identifying those indirect studies, which was to be expected as these articles do not answer our focused PICO question. We therefore opted to maintain the initial search strategy, combining the administrative databases and definitions/algorithms terms with an OR (the most sensitive search strategy).

MEDLINE Search 11h November 2024

| | Combined terms | Outputs |
|---|---|---|
| **#1** | "algorithm*"[MeSH Terms] OR "Current Procedural Terminology"[MeSH Terms] OR "International Classification of Diseases"[MeSH Terms] OR "diagnostic algorithm*"[Text Word] OR "classification algorithm*"[Text Word] OR "claim based algorithm*"[Text Word] OR "Diagnostic codes"[Text Word] OR "diagnostic definition*"[Text Word] OR "ICD codes"[Text Word] OR "ICD"[Text Word] OR "ACR Criteria"[Text Word] | 549,088 |
| **#2** | "databases, factual"[MeSH Terms] OR "Database Management Systems"[MeSH Terms] OR "medical records systems, computerized"[MeSH Terms] OR "Electronic Health Records"[MeSH Terms] OR "Medical Record Linkage"[MeSH Terms] OR "administrative claims, healthcare"[MeSH Terms] OR "Hospital Records"[MeSH Terms] OR "Patient Discharge"[MeSH Terms] OR "Electronic Data Processing"[MeSH Terms] OR "Automatic Data Processing"[Text Word] OR "Insurance database"[Text Word] OR "Record linkage"[Text Word] OR "Data Warehouse"[Text Word] OR "Registered Person Database"[Title/Abstract:~3] OR "Electronic Medical Records"[Text Word] OR ("Billing"[All Fields] AND ("Data"[All Fields] OR "Claims"[All Fields] OR "Codes"[All Fields])) OR ("Claims"[All Fields] AND ("Administrative"[All Fields] OR "data*"[All Fields])) OR (("Administrative"[All Fields] OR "Healthcare"[All Fields]) AND ("data*"[All Fields] OR "Records"[All Fields])) OR "administrative data*"[Text Word] OR "Administration Database"[Text Word] OR "Physician claims"[Text Word] OR "Hospital Billing"[Text Word] | 706,807 |
| **#3** | ("Validation Study"[Publication Type] AND "Sensitivity and Specificity"[MeSH Terms]) OR "Predictive Value of Tests"[MeSH Terms] OR "Reproducibility of Results"[MeSH Terms] OR "Data Accuracy"[MeSH Terms] OR "ROC Curve"[MeSH Terms] OR "Receiver Operating Characteristic | 3,169,224 |

| | | |
|---|---|---|
| | Curve"[Title/Abstract:~4] OR "Predictive value"[Text Word] OR "Diagnostic Accuracy"[Title/Abstract:~3] OR "Positive Predictive Value"[Title/Abstract:~3] OR "Negative Predictive Value"[Title/Abstract:~3] OR "Accuracy"[Text Word] OR "Sensitivity"[Text Word] OR "Specificity"[Text Word] OR "Validity"[Text Word] OR "Validation Process"[Text Word] OR "Case Ascertainment"[Text Word] OR "Diagnostic performance"[Text Word] | |
| **#4** | (#1 OR #2) AND #3 | 251,729 |
| **#5** | "Comment"[Publication Type] OR "Case Reports"[Publication Type] OR "Editorial"[Publication Type] OR "Literature Review"[All Fields] | 4,072,939 |
| **#6** | #4 NOT #5 | 248,230 |
| **#7** | "Arthritis, Rheumatoid" [Mesh] OR "Arthritis, Juvenile" [Mesh] OR "Felty Syndrome" [Mesh] OR "Caplan Syndrome" [Mesh] OR (("Arthritis, Rheumatoid"[Mesh]) AND "Lung Diseases, Interstitial"[Mesh]) OR "Rheumatoid Arthritis" [Text word] OR "Rheumatoid Vasculitis" [Text word] OR "Rheumatoid Nodule" [Text word] OR "Rheumatoid Nodules" [Text word] OR "Juvenile Idiopathic Arthritis" [Title/Abstract: ~3] OR "Juvenile Chronic Arthritis" [Title/Abstract: ~3] OR "Juvenile Rheumatoid Arthritis" [Title/Abstract: ~3] OR "Felty Syndrome" [Text word] OR "Caplan Syndrome" [Text word] OR (("Rheumatoid Arthritis") AND ("Interstitial Lung Disease")) OR "RA-ILD" [Text word] OR "(rheumat* AND arthritis)" | 184,236 |
| **#8** | #6 AND #7 | 1,030 |

CENTRAL Search 11th November 2024

| ID | Search Hits | Outputs |
|---|---|---|
| **#1** | [mh "algorithm"] | 8223 |
| **#2** | [mh "algorithms"] | 8223 |
| **#3** | [mh "Current Procedural Terminology"] | 5 |
| **#4** | [mh "International Classification of Diseases"] | 117 |
| **#5** | diagnostic NEXT algorithm*:ti,ab,kw | 228 |
| **#6** | classification NEXT algorithm*: ti,ab,kw | 185 |
| **#7** | claim NEXT based NEXT algorithm*:ti,ab,kw | 4 |
| **#8** | "Diagnostic codes":ti,ab,kw | 90 |
| **#9** | Diagnostic NEXT Definition*:ti,ab,kw | 11 |
| **#10** | "ICD codes":ti,ab,kw | 66 |
| **#11** | "ICD":ti,ab,kw | 6328 |
| **#12** | "ACR Criteria":ti,ab,kw | 1271 |
| **#13** | #1 OR #2 OR #3 OR #4 OR #5 OR #6 OR #7 OR #8 OR #9 OR #10 OR #11 OR #12 | 16165 |

| | | |
|---|---|---|
| #14 | [mh "databases, factual"] | 1411 |
| #15 | [mh "Database Management Systems"] | 34 |
| #16 | [mh "medical records systems, computerized"] | 1218 |
| #17 | [mh "Electronic Health Records"] | 898 |
| #18 | [mh "Medical Record Linkage"] | 40 |
| #19 | [mh "administrative claims, healthcare"] | 13 |
| #20 | [mh "Hospital Records"] | 18 |
| #21 | [mh "Patient Discharge"] | 2767 |
| #22 | [mh "Electronic Data Processing"] | 136 |
| #23 | "Automatic Data Processing":ti,ab,kw | 3 |
| #24 | "Insurance database":ti,ab,kw | 88 |
| #25 | "Record linkage":ti,ab,kw | 161 |
| #26 | "Data Warehouse":ti,ab,kw | 170 |
| #27 | "Registered Person Database":ti,ab,kw | 0 |
| #28 | "Electronic Medical Records":ti,ab,kw | 1154 |
| #29 | (("Billing") AND ("data" OR "claims" OR "codes")):ti,ab,kw | 329 |
| #30 | (("Claims") AND ("Administrative" OR "Data")):ti,ab,kw | 1804 |
| #31 | Administrative NEXT data*:ti,ab,kw | 1010 |
| #32 | "Administration Database":ti,ab,kw | 10 |
| #33 | "Physician claims":ti,ab,kw | 12 |
| #34 | "Hospital Billing":ti,ab,kw | 63 |
| #35 | ("Claims" AND ("Administrative" OR "data")):ti,ab,kw | 1804 |
| #36 | (("Administrative" OR "Healthcare") AND ("Data" OR "Records")):ti,ab,kw | 18006 |
| #37 | #14 OR #15 OR #16 OR #17 OR #18 OR #19 OR #20 OR #21 OR #22 OR #23 OR #24 OR #25 OR #26 OR #27 OR #28 OR #29 OR #30 OR #31 OR #32 OR #33 OR #34 OR #35 OR #36 | 25819 |
| #38 | #13 OR #37 | 41151 |
| #39 | [mh "Sensitivity and Specificity"] | 21953 |
| #40 | [mh "Predictive Value of Tests"] | 10133 |
| #41 | [mh "Reproducibility of Results"] | 16684 |
| #42 | [mh "Data Accuracy"] | 152 |
| #43 | [mh "ROC Curve"] | 2187 |
| #44 | "Receiver Operating Characteristic Curve":ti,ab,kw | 1751 |
| #45 | "Predictive value":ti,ab,kw | 17907 |
| #46 | Diagnostic near/3 Accuracy:ti,ab,kw | 9715 |
| #47 | "Positive Predictive Value":ti,ab,kw | 2634 |
| #48 | "Negative Predictive Value":ti,ab,kw | 2236 |
| #49 | "Accuracy":ti,ab,kw | 30828 |
| #50 | "Sensitivity":ti,ab,kw | 75359 |
| #51 | "Specificity":ti,ab,kw | 25214 |
| #52 | "Validity":ti,ab,kw | 14676 |
| #53 | "Validation Process":ti,ab,kw | 4189 |
| #54 | "Case Ascertainment":ti,ab,kw | 36 |
| #55 | "Diagnostic performance":ti,ab,kw | 1315 |
| #56 | #39 OR #40 OR #41 OR #42 OR #43 OR #44 OR #45 OR #46 OR #47 OR #48 OR #49 OR #50 OR #51 OR #52 OR #53 OR #54 OR #55 # | 135226 |
| #57 | #38 AND #56 | 6707 |
| #58 | (“Comment”):pt OR (“Case Reports”):pt OR (“Editorial”):pt OR (“Literature Review”) | 2519 |

| | | |
|---|---|---|
| #59 | #57 NOT #58 | 6643 |
| #60 | [mh "Arthritis, Rheumatoid"] | 8149 |
| #61 | [mh "Arthritis, Juvenile"] | 464 |
| #62 | [mh "Felty Syndrome"] | 0 |
| #63 | [mh "Caplan Syndrome"] | 0 |
| #64 | [mh "Arthritis, Rheumatoid"] AND [mh "Lung Diseases, Interstitial"] | 23 |
| #65 | "Rheumatoid Arthritis":ti,ab,kw | 18690 |
| #66 | "Rheumatoid Vasculitis":ti,ab,kw | 7 |
| #67 | "Rheumatoid Nodule":ti,ab,kw | 17 |
| #68 | "Rheumatoid Nodules":ti,ab,kw | 38 |
| #69 | "Juvenile Idiopathic Arthritis":ti,ab,kw | 795 |
| #70 | "Juvenile Chronic Arthritis":ti,ab,kw | 53 |
| #71 | "Juvenile Rheumatoid Arthritis":ti,ab,kw | 573 |
| #72 | "Felty Syndrome":ti,ab,kw | 6 |
| #73 | "Caplan Syndrome":ti,ab,kw | 0 |
| #74 | (("Rheumatoid Arthritis") AND ("Interstitial Lung Disease")):ti,ab,kw | 86 |
| #75 | "RA ILD":ti,ab,kw | 30 |
| #76 | rheumat* AND arthritis:ti,ab,kw | 23152 |
| #77 | #60 OR #61 OR #62 OR #63 OR #64 OR #65 OR #66 OR #67 OR #68 OR #69 OR #70 OR #71 OR #72 OR #73 OR #74 OR #75 OR #76 | 23774 |
| #78 | #59 AND #77 | 93 |

Embase Search 12th November 2024

| | | |
|---|---|---|
| #1 | 'algorithm'/exp OR 'algorithm' OR 'current procedural terminology'/exp OR 'current procedural terminology' OR 'international classification of diseases'/exp OR 'international classification of diseases' OR 'diagnostic algorithm*':ti,ab,kw OR 'classification algorithm':ti,ab,kw OR 'claims based algorithm':ti,ab,kw OR 'diagnostic codes':ti,ab,kw OR 'diagnostic definition':ti,ab,kw OR 'icd codes':ti,ab,kw OR 'icd':ti,ab,kw OR 'acr criteria':ti,ab,kw | 965,982 |
| #2 | 'factual database'/exp OR 'factual database' OR 'database management system'/exp OR 'database management system' OR 'electronic medical record system'/exp OR 'electronic medical record system' OR 'electronic health record'/exp OR 'electronic health record' OR 'medical record'/exp OR 'medical record' OR 'administrative claims (health care)'/exp OR 'administrative claims (health care)' OR 'hospital discharge'/exp OR 'hospital discharge' OR 'insurance database':ti,ab,kw OR 'record linkage':ti,ab,kw OR 'data warehouse':ti,ab,kw OR 'registered next/2 person next/2 database':ti,ab,kw OR 'electronic medical records':ti,ab OR ('billing':ti,ab,kw AND ('data':ti,ab,kw OR 'claims':ti,ab,kw OR 'codes':ti,ab,kw)) OR ('claims':ti,ab,kw AND ('administrative':ti,ab,kw OR 'data*':ti,ab,kw)) OR (('administrative':ti,ab,kw OR 'healthcare':ti,ab,kw) AND ('data*':ti,ab,kw OR 'records':ti,ab,kw)) OR 'administrative data*':ti,ab,kw OR 'administration database':ti,ab,kw OR 'physician claims':ti,ab,kw OR 'hospital billing':ti,ab,kw | 1,230,052 |
| #3 | #1 OR #2 | 2,089,508 |
| #4 | 'sensitivity and specificity'/exp OR 'sensitivity and specificity' OR 'predictive value'/exp OR 'predictive value' OR 'reproducibility'/exp OR 'reproducibility' OR 'data accuracy'/exp OR 'data accuracy' OR 'receiver | 3,213,131 |

| | | |
|---|---|---|
| | operating characteristic'/exp OR 'receiver operating characteristic' OR 'receiver operating characteristic curve':ti,ab,kw OR 'predictive value':ti,ab,kw OR 'diagnostic next/2 accuracy':ti,ab,kw OR 'positive next/2 predictive value':ti,ab,kw OR 'negative next/2 predictive value':ti,ab,kw OR 'accuracy':ti,ab,kw OR 'sensitivity':ti,ab,kw OR 'specificity':ti,ab,kw OR 'validity':ti,ab,kw OR 'validation process':ti,ab,kw OR 'case ascertainment':ti,ab,kw OR 'diagnostic performance':ti,ab,kw | |
| **#5** | #3 AND #4 | 363,956 |
| **#6** | 'comment':it OR 'case reports':it OR 'editorial':it OR 'literature review':it | 812,392 |
| **#7** | #5 NOT #6 | 362,739 |
| **#8** | 'rheumatoid arthritis'/mj OR 'rheumatoid arthritis' OR (('rheumatoid arthritis'/mj OR 'rheumatoid arthritis') AND ('interstitial lung disease'/mj OR 'interstitial lung disease')) OR 'rheumatoid arthritis':ti,ab,kw OR 'rheumatoid vasculitis':ti,ab,kw OR 'caplan syndrome':ti,ab,kw OR ('rheumatoid arthritis':ti,ab,kw AND 'interstitial lung disease':ti,ab,kw) OR 'ra-ild':ti,ab,kw OR ('rheumat*':ti,ab,kw AND 'arthritis':ti,ab,kw) | 307,962 |
| **#9** | #7 AND #8 | 2,987 |

### Supplementary Appendix 3. Methods

Data items

The target condition was a diagnosis of RA recorded in EHRs or other administrative databases. The “index” classification was a definition (codes) or algorithm used to identify RA in EHRs and the “reference standard” the register of a diagnosis for RA in medical records or confirmed by a rheumatologist, or any other reference standard as defined by the authors of the original study.

True positives (TP) cases were considered as a diagnosis of RA by a code or algorithm confirmed by the “reference standard” (i.e., medical records review) and true negatives (TN) as cases not identified with RA by both definition or algorithm and the “reference standard”. False positives (FP) were considered as those cases identified with RA by a definition or algorithm in EHRs but not confirmed by the “reference standard” and false negatives (FN) as those cases not identified by the definition or algorithm in the EHRs but confirmed with RA by the “reference standard”.

Diagnostic accuracy measures

Primary diagnostic accuracy measures were sensitivity, specificity, positive predictive value (PPV) and negative predictive value (NPV). When a study did not provide results on these measures, we used the data from the two-by-two tables to calculate the accuracy statistics when available. For a practical interpretation of the accuracy measures, studies with values ≥80% were considered to have high accuracy, values from 60-80% moderate accuracy and values <60% were considered to have low accuracy as previously reported. The unit of assessment was at the individual level. The statistics were calculated following the formulas:

| | |
|---|---|
| Sensitivity: | TP / (TP + FN) |
| Specificity: | TN / (TN + FP) |
| PPV: | TP / (TP + FP) |
| NPV: | TN / (TN + FN) |

Synthesis of results

We classified the definitions and algorithms in the studies according to the number and type of codes (e.g., ICD, prescriptions, diagnostic test, procedural codes) used to ascertain the diagnosis of RA. Additionally, we were guided by the classification used by Shrestha et al [18] in their systematic review to classify the algorithms (with modifications made as necessary):

- *Less restrictive algorithms*: those that required a single diagnostic code for the condition under study from one outpatient visit or unspecified source (i.e., single ICD code for RA). An algorithm requiring a single diagnostic code from one outpatient visit or one prescription record was classified as less restrictive as the stricter code (prescription) is not required to identify the condition (as the definition is based on presence of either code).

- *Restrictive algorithms*: those that required more than one code of any kind or if it required one or more strict code such a procedural, prescription, diagnostic test, or hospitalisation codes (rather than a single diagnosis/disease code). For example:
  - An algorithm that required a diagnosis ICD code from two separate outpatient visits.
  - An algorithm that required one ICD code from an outpatient visit and one prescription code.
  - An algorithm that required a single hospitalisation visit.

The definitions and algorithms were generally classified as restrictive or less restrictive as described above. Furthermore, we developed sub-categories among restrictive algorithms to stratify analysis, based on anticipated accuracy and how frequently they were evaluated by included studies. We identified 4 main sub-categories:

- <u>RA diagnoses codes + DMARD prescription codes</u>: These algorithms require at least the presence of ICD codes and DMARD prescriptions to identify RA cases. Some algorithms are based solely on this combination of codes, while others include more specific prescription codes (i.e., biologic agents, glucocorticoids) in their algorithms and ≥1 ICD code for RA. For example:
  a. >2 ICD codes for RA + prescription codes (DMARDs).
  b. 1 ICD code for RA (hospitalisation) OR (≥2 ICD codes for RA + DMARDs OR biologic agents).
- <u>≥2 ICD codes for RA:</u> These algorithms require at least 2 ICD codes for RA in EHRs on different dates, without combination with other types of codes (i.e., prescription codes,

laboratory values). This only includes non-specific coding, and requirements for diagnoses to be recorded by a rheumatologist or specialist are included in sub-category 3. For example:

a. ≥2 ICD codes for RA by physician

b. ≥2 ICD codes recorded in claims data

- ≥1 ICD code for RA by rheumatologist: These algorithms include and are limited to ≥1 ICD codes for RA recorded by a rheumatologist or specialist with or without other ICD codes for RA (i.e., physician or claims data) and without combination with other types of codes (i.e., prescription codes or laboratory values). For example:

a. ≥2 ICD codes for RA by rheumatologist

b. 2 RA codes (physician) with ≥1 by specialist in 1 year

- ≥1 ICD hospitalisation code for RA: These algorithms require at least one ICD hospitalisation code for identifying RA in EHRs. They may or may not be combined with other ICD codes for RA (i.e., outpatient, physician or emergency ICD codes). For example:

a. 1 RA code ever (hospitalisation)

b. 1 RA code (hospitalisation) OR ≥2 RA codes (outpatient)

### Supplementary Appendix 4. Table 1

Table 1. Characteristics of included studies (n= 35)

| Author, Year, Country | Study Design | Administrative Data Source, Time period | Record type | Study Population N | Sample size | Gender | Age (years) | Diagnosis | Codes for Algorithm definition | Reference standard |
|---|---|---|---|---|---|---|---|---|---|---|
| Allebeck et al. 1983, Sweden | Cross-sectional study | Stockholm county Medical Information System<br><br>1975-1978 | Hospital Discharge Records | Female patients between 15-50 years with a sole diagnosis of RA<br><br>N= NR | n= 291<br><br>analysed= 276 | F= 100% | 15-50 | RA | ICD-8 codes for RA (712.38, 712.39) | Medical record review by one author based on diagnostic criteria for RA from Rome (1961) and New York (1966) |
| Almutairi et al. 2021, Australia | Retrospective Cohort study | Finance and Business Office at Sir Charles Gairdner Hospital (SCGH), Perth<br><br>2008-2020 | Hospital Discharge Records | Patients listed in the SCGH AHD with a primary or secondary ICD 10-AM diagnostic discharge record for RA<br><br>N= NR | n=200<br><br>Analysed= 87 | F= 67.8% | 64.7 ± 17.2 | RA | ICD-10-AM codes for RA (M05.0-M06.9)<br><br>Biological infusion AR-DRG code I40Z | Medical charts from the Medical Records Department at SCGH (Rheumatologist-reported diagnosis and ACR/EULAR classification) |
| Carrara et al. 2015, Italy | Nested Case-Control and Cohort diagnostic study | *Training set:*<br>Rheumatology Unit, IRCCS Policlinico San Matteo Foundation, Pavia<br>2007-2010<br><br>*Validation set:*<br>Secondary rheumatology centre: EMRs of the Rheumatology outpatient clinic of the Clinical Institute Beato Matteo of Vigevano<br><br>Primary care EMRs: convenience sample of six primary care physicians of the Local Health Authority of Pavia | NHS assistance (demographic and administrative records)<br><br>Certification of CDs for the exemption from co-payment<br><br>Hospital Discharge Records (HDRs)<br><br>Outpatient Drug Prescription Records | Patients ≥16 years assisted by a tertiary rheumatology clinic (Rheumatology Unit, IRCCS Policlinico San Matteo Foundation)<br><br>N= NR | *Training set:*<br>n=900<br>Analysed= 827<br>RA= 301<br>Non-RA= 526<br><br>*Validation set:*<br>n= 138<br>Analysed= 106<br>RA= 32<br><br>n=6087<br>Analysed= 6087 | RA=<br>F= 72.4%<br><br>Non-RA=<br>F= 77% | RA=<br>66.8 ± 13.1<br><br>Non-RA=<br>57.7± 15.7 | RA | RA certification by rheumatologist/absence of certification of other CADs.<br><br>ICD-9-CM code 714.0 in HDRs<br><br>DMARDs prescriptions | Medical Records clinically validated by an external investigator or according to specific classification criteria |

| | | | | | | | | | | |
|---|---|---|---|---|---|---|---|---|---|---|
| | | | | | RA= 40 | | | | | |
| Carroll et al.^<br>2012<br>USA | Retrospective Cohort study | Vanderbilt University Medical Center's Synthetic Derivative (VUMC SD)<br><br>Northwestern medical Enterprise Data Warehouse (EDW)<br><br>Partners Healthcare (PH) database used by BWH and MGH<br><br>NR | Electronic Health Records (linked to BioVU repository)<br><br>Electronic Health Records (Hospital Records)<br><br>Hospital Records<br><br>Narrative EMRs data | VUMC SD:<br>≥ 18 adults accrued into BioVU with ≥1 ICD-9 code for RA (714.*)<br>N= 10,000<br><br>EDW:<br>Patients with ≥1 ICD-9 code for RA (714.*)<br>N= 6,124<br><br>PH:<br>Patients with ≥1 ICD-9 code for RA (714.*)<br>N= 29,432 | VUMC SD<br>n= 376<br><br>EDW<br>n= 400<br>PH:<br>n= 500 | VUMC SD<br>RA=<br>F= 80%<br>Non-RA=<br>F= 73.8%<br>EDW<br>RA=<br>F= 81.4%<br>Non-RA=<br>F= 72.6%<br><br>PH<br>RA=<br>F= 77.1%<br>Non-RA=<br>75% | VUMC SD<br>RA=<br>52.9 ± 13.1<br>Non-RA=<br>56.2 ± 16.5<br>EDW<br>RA=<br>54.3 ± 14.8<br>Non-RA=<br>58.9 ± 16.8<br><br>PH:<br>RA=<br>60.7 ± 15.9<br>Non-RA=<br>56.0 ± 18.6 | RA | >1 ICD-9 RA code (714.xx)<br><br>Medication prescriptions<br><br>RF and anti-CCP lab results | Medical chart review by rheumatologists |
| Cho et al.<br>2013,<br>South Korea | Retrospective Cohort study | The Korean National Health Insurance (NHI) claims database<br><br>July 2009-December 2009 | Administrative and Claims data | Patients with diagnostic code for seropositive RA (M05.xx) when individual co-payment beneficiaries program began (July 2009)<br><br>N= 73,858 | n= 59,823<br><br>RA=<br>n= 50,082<br><br>Non-RA=<br>9,741 | F= 80.3% | 55.8 ± 12.9 | Seropositive RA | ICD-10 code for RA (M05.xx)<br><br>Drug prescriptions | Official report from a doctor documenting that the patient fulfils the 1987 ACR criteria (registered in the program) |
| Convertino et al.<br>2021,<br>Italy | Retrospective Cohort study | Tuscan Healthcare Administrative Databases (THAD)<br>2014-2016 | Hospital discharge records<br><br>Emergency Department Accesses records<br><br>Drugs supply and exemption co-payments | First-ever users of DMARDs with at least one record of visit to the Rheumatology Unit<br><br>N= NR | n=277 | NR | 53.3 ± 13.9 | RA | ICD-9 codes for RA (714*)<br><br>Exemption code from co-payment (006) | Chart Medical Records of the Rheumatology Unit of Pisa University Hospital; diagnosis by rheumatologist |

| | | | | | | | | | | |
|---|---|---|---|---|---|---|---|---|---|---|
| Curtis et al. 2018, USA | Retrospective Cohort study | Administrative Medical and pharmacy claims data from Medicare and a commercial Health Plan<br><br>2005-2012 | Claims data | RA patients participating in the Corrona (North America RA registry) linked to Medicare/Commercial health plan<br><br>N= NR | n= 6,509 | Medicare F=74.8%<br><br>Commercial Claims F= 80.1% | Medicare 67.2 ± 9.8<br><br>Commercial Claims= 48.1 ± 10.6 | Incident RA | ICD-9 codes for RA (714.0, 714.2, 714.81, 714.x)<br><br>Medications | Clinical diagnosis by rheumatologist in the Corrona registry |
| Fowles et al. 1995, USA | Retrospective Cohort study | Medicare Part B<br><br>1990-1991 | Claims data | Maryland Medicare beneficiaries<br><br>N= 1,998 | n= 1,998<br><br>Analysed= 1,596 | NR | NR | RA | ICD-9-CM codes for RA | Medical record review by trained nurses |
| Hanly et al. 2015, Canada | Matched Case-Control study | Medical Services Insurance (MSI) program, Nova Scotia Health Data Nova Scotia databases (3)<br><br>1997-2011 | Claims data<br><br>Hospital Discharge records<br><br>Physician billings | Nova Scotia residents enrolled in the MSI program with ≥1 rheumatologist consultation at the Arthritis Center of Nova Scotia<br><br>N= 25,888 (RA= 2,692) | RA= n= 535<br><br>non-RA= n= 2,140 | F= 67.9% | 56.0 ± 17.8 | Prevalent and Incident RA | ICD-9 and ICD-10 codes for RA (714.0, 714.1, 714.2, M05-M05.9, M06.0, M06.8, M06.9) | Medical chart review Rheumatologist's diagnosis at the Arthritis Center of Nova Scotia |
| Harrold et al. 2013, USA | Retrospective Cohort study | Kaiser Permanente, Northern California (KPNC) Autoimmune Disease Registry<br><br>1996-2009 | Electronic Medical Records | Children ≤15 years with ≥1 ICD-9 code for JIA (696.0, 714, 720) enrolled in the KPNC<br><br>N= 1,153 | n= 97<br><br>JIA cases= 67 | JIA cases= F= 64% | JIA cases= 6-10= 21% 11-15= 46% | Incident and Prevalent JIA | ICD-9 codes (696.0, 714, 720)<br><br>Laboratory tests | Manual chart review (Clinical diagnosis by adult or pediatric rheumatologist) |
| Huang et al.^ 2020, USA | Retrospective Cohort study | EMRs data from two large academic hospitals in Boston (BWH and MGH)<br><br>1994/1996-2017 | Hospital discharge records<br><br>Claims data<br><br>Narrative EMRs data | Patients with ≥1 RA ICD-9 or ICD-10 code and ≥2 notes with length >500 characters in EMRs (RA data mart)<br><br>N= 53,144 | n=200<br><br>n=100 (RA ICD-10 codes only) | RA= F= 78.7%<br><br>Non-RA= F= 71.2% | RA= 65.4 ± 16.1<br><br>Non-RA= 66.4 ± 17.4 | RA | ICD-9 and ICD-10 RA codes (714.x, M05.x, M06.x)<br><br>RF and anti-CCP lab results<br><br>DMARDs prescriptions | Medical chart review, RA diagnosis by a rheumatologist Presence of 2010 ACR/EULAR criteria |

| | | | | | | | | | | |
|---|---|---|---|---|---|---|---|---|---|---|
| Ibfelt et al. 2017, Denmark | Retrospective Cohort study | The Danish Rheumatologic Database (DANBIO)<br><br>The Danish National Patient Registry (DNPR)<br><br>2001-2011 | Clinical and treatment data by rheumatologists<br><br>Hospital records | Patients registered with an RA diagnosis in DANBIO and DNRP in 2011<br><br>N= 2,298 | n=1,678<br><br>Analysed= n=1,532 | NR | NR | Incident RA | ICD-10 RA diagnostic codes (M05.9, M06.0, M06.8, M06.9) | Medical record review by rheumatologist diagnosis |
| Katz et al. 1997, USA | Cross-sectional study | 8 Rheumatology practices in Massachusetts, Colorado, and Virginia<br>Medicare (Part B)<br><br>03/1993-10/1993 | Physician claims (Part B) | Patients with RA documented in the medical records at the 8 rheumatology practices<br><br>N= NR | n= 160<br>Analysed: 153 | NR | NR | RA | ICD-9-CM code for RA (714.0, 714.1, 714.2, 714.3, 714.30, 714.31, 714.32, 714.33)<br><br>CPT codes (20550, 20600, 20605, 20610) | Medical records review, diagnosis by rheumatologists and 1990 ACR criteria |
| Kim et al. 2011, USA | Retrospective Cohort study | Medicare and the Pennsylvania Assistance Contract for the Elderly (PACE) program<br><br>1994-2004 | Claims data | Pennsylvania residents ≥65 years, low-moderate income<br><br>N= 9,482 | n= 158 | F= 82.9% | 79.3 ± 7.1 | RA | ICD-9 code for RA (714)<br><br>DMARDs prescriptions | Medical records by rheumatologists<br><br>Definition: diagnosis of RA by rheumatologist and fulfilment of 1987 ACR criteria |
| Kronzer et al.[¥] 2020, USA | Retrospective Cohort study | Mayo Clinic Biobank included in the Rochester Epidemiology Project (REP)<br><br>1995-2009 | Electronic Health Records | Mayo Clinic Biobank participants included in the REP with ≥1 ICD-9 RA code<br><br>N= NR | n= 497<br><br>RA= 213<br><br>Non-RA= 284 | RA= F= 75%<br><br>Non-RA= F= 68% | RA= 65 ± 14<br><br>Non-RA= 63 ± 16 | Prevalent and Incident RA | eMERGE algorithm:<br><br>ICD-9 code for RA (714.x)<br><br>RF lab value<br><br>ICD-9 codes for SLE and PA *(exclusion criteria)* | Manual chart review against 1987 ACR or 2010 ACR/EULAR criteria |
| Kubota et al. 2021, Japan | Retrospective Cohort study | 64 Hospitals of Tokushukai Medical Group, Tokushukai Information System | Claims data<br><br>Clinical data | Patients with a RA code in at least 1 monthly claim | n= 19,734<br><br>Analysed= | F= 50.3% | ≤24 yr= 17.7% | RA | ICD-10 codes for RA | Chart review by rheumatologists |

| | | | | | | | | | | |
|---|---|---|---|---|---|---|---|---|---|---|
| | | 2018-2019 | | N= 1,590,669 | 12,982 | | 25-64 yr= 42,8%<br><br>≥65 yr= 39,5% | | Medication prescription<br><br>ICD-10 codes for SADs *(exclusion criteria)* | |
| Liao et al.^<br>2010,<br>USA | Retrospective Cohort study | EMRs data from two large academic hospitals in Boston (BWH and MGH)<br><br>1994/1996-2008 | Hospital discharge records<br><br>Claims data<br><br>Narrative EMRs data | Patients with >1 ICD-9 RA code (714.xx) or anti-CCP testing (RA data mart)<br><br>N= 29,432 | Training set: n= 500<br><br>Validation set: n= 400 | Training set RA= F= 77%<br><br>Non-RA F= 74% | Training set RA= 60.4 ± 16<br><br>Non-RA= 56.1 ± 19 | RA | >1 ICD-9 RA code (714.xx)<br><br>Electronic prescriptions<br><br>RF and anti-CCP lab results | Medical record review by blinded 2 rheumatologists (clinical diagnosis and 1987 ACR criteria) |
| Linauskas et al. 2018,<br>Denmark | Retrospective cohort study | The Danish Diet, Cancer, and Health Cohort study<br><br>The Danish National Patient Registry (DNPR)<br><br>The Danish National Prescription Registry<br><br>1977-2016 | Hospital Discharge records<br><br>Drug reimbursement data | Participants from the Danish Diet, Cancer, and Health Cohort with ≥1 RA (712.39, M05, M06) code registered at hospitals in the DNPR<br><br>N= 331 | n= 331 | F= 70% | 65<br>(35-83) | Incident RA | ICD-8 – ICD-10 codes for RA (712.39, M05, M06)<br><br>DMARDs prescription | Medical records, RA diagnosis verified against 1958 ACR, 1987 ACR or 2010 ACR/EULAR criteria. If unmet, clinical assessment by rheumatologist |
| Losina et al. 2003,<br>USA | Cross-sectional study | Medicare database Ohio, Pennsylvania, and Colorado<br><br>1995 | Claims data (hospital and surgeon's) | Inpatient Medicare beneficiaries who received primary THR in 1995<br><br>N= NR | n= 922 | NR | NR | RA | ICD-9-CM for RA (714, 714.0) | Medical record review from Physician Review Organizations in each state by trained nurses |
| Nanji et al 2012,<br>Canada | Retrospective Cohort study | 2 primary care clinics in Grande Prairie, Alberta (4 family physicians)<br><br>NR | Electronic Health Records | Patients with RA codes in the EHRs of 4 family physicians<br><br>N= NR | n= 31 | NR | NR | RA | ICD-9 codes for RA (714, 714.0) | Chart review by rheumatologist clinical diagnosis |
| Ng et al. 2012,<br>USA | Retrospective Cohort study | The Veterans Health Administration (VHA) database-Michael E. DeBakey VAMC, Houston | Electronic Health records<br><br>Claims data | Patients with ≥2 ICD-9 RA codes at least 6 months apart | n= 543 | F= 9% | 62 ± NR | RA | ≥2 ICD-9 codes for RA (714) at least 6 months apart | Medical record review<br><br>2 ICD-9 codes as noted + at least 4 1987 ACR criteria or |

| | | | | | | | | | | |
|---|---|---|---|---|---|---|---|---|---|---|
| | | 1998-2009 | | N= 1,779 | | | | | DMARDs prescriptions | positive anti-CCP or patient self-report of RA management by non-VA rheumatologist |
| Paltta et al. 2021, Finland | Case-Control study | Five Hospital Biobanks in Finland<br>Finnish Care Register for Health Care (CRHC)<br><br>Finnish National Health Insurance system<br><br>2007-2018 | Hospital Discharge records<br><br>Medical reimbursement registers for DMARDs | Patients from 5 Hospital Biobanks in Finland<br><br>N= NR | RA= 250 Analysed= 233<br><br>Non-RA= 250 | F= 69% | Me: 50 (IQR 40-59) at diagnosis | Seropositi ve and seronegati ve RA | ICD-10 codes for RA (M0.58, M05.9 and M06.0)<br><br>DMARDs reimbursemen t codes 202, 281, 313 for RA | Chart review by a rheumatologist or resident in rheumatology |
| Pedersen et al. 2004, Denmark | Retrospective Cohort study | The Danish Diet, Cancer, and Health Cohort study and the Danish Nationwide Twin Population linked to The Danish National Patient Registry (DNPR)<br><br>1977-2001 | Hospital Discharge records | Patients >15 years registered in the DNPR with ICD-8 or ICD-10 code for RA from the 2 cohorts<br><br>N= 48,707 | n= 217 | F : M ratio 2.7 : 1 | 57 (15-72) | RA | ICD-8, ICD-10 codes for RA (712.19, 712.39, 712.59, M05, M06) | Medical records review by rheumatologist (clinically confirmed RA and 1987 ACR criteria) |
| Peterson et al. 2023, USA | Retrospective Cohort study | Vanderbilt University Medical Center (VUMC) database<br><br>1990-NR | Electronic Health records (clinical, laboratory, medication data) | ≥1 ICD-9 or ICD-10 codes for JIA<br><br>N= 3,190 | Training set n= 200<br><br>Validation set n= 100 | JIA= F= 75%<br><br>Non-JIA= F= 63% | JIA= Median= 19 (13-23)<br><br>Non-JIA= Median= 32 (22-48) | JIA | ICD-9, (714.3, 714.30, 714.31, 714.32, 714.33) ICD-10 (M08.0, M08.1-M08.4, M08.8, M08.9, L40.54) codes for JIA | Chart review by paediatric rheumatologist (JIA ICD code at age ≤20 years and a rheumatology clinic note documenting diagnosis) |
| Singh et al. 2004, USA | Retrospective Cohort study | Minneapolis Veterans Affair (VA) administrative health database (AHDs)<br><br>2001- 2002 | Electronic Health records (administrative, pharmacy and laboratory records) | Patients at the Minneapolis VA rheumatology clinic<br><br>N= 737 | n= 252 Analysed= 184 | F= 6% | 64.4 ± 12.9 | RA | ICD-10 code 714.0 for RA<br><br>DMARDs prescriptions | Chart review from medical records: RA diagnosis by a rheumatologist on 2 separate occasions >6 weeks apart |

| | | | | | | | | | | |
|---|---|---|---|---|---|---|---|---|---|---|
| | | | | | | | | | Positive RF code | |
| Shiff et al.<br>2017,<br>Canada | Retrospective Cohort study | The Population Health Research Data Repository: Manitoba health insurance registry<br><br>Hospital Discharge Abstracts Database (DAD)<br><br>Medical Claims Database<br><br>1980-2012 | Insurance coverage data<br><br>Hospital Discharge records<br><br>Physician billings | Children <16 yrs seen by pediatric rheumatologist at the Children's Hospital, Winnipeg, with a diagnosis of rheumatic disease from the Pediatric Rheumatology Clinical Database<br><br>N= 1,812 | n= 1,122<br><br>JIA= 707<br><br>Non-JIA= 415 | JIA=<br>F= 68.7%<br><br>Non-JIA=<br>F= 64.6% | JIA=<br>6-10= 21%<br>11-15= 44%<br><br>Non-JIA=<br>6-10= 24%<br>11-15= 46.5% | JIA | ICD-9-CM (714, 720), ICD-10-CA (M05, M06, M08, M45) codes for JIA | Diagnosis by a pediatric rheumatologist |
| Stringer et al.<br>2015,<br>Canada | Cross-sectional study | Nova Scotia (NS) Medical Service Insurance (MSI) database<br><br>Clinical database of the pediatric rheumatology clinic, IWK Health Centre<br><br>2005/2006-2008/2009 | Insurance data | Children <16 years with ICD-9 code for JIA in NS<br><br>N= NR | n= 94 | NR | NR | Incident JIA | ICD-9 code for JIA (714) | Clinical chart review (diagnosis by rheumatologist based on 2004 ILAR criteria) |
| Sugiyama et al.<br>2024,<br>Japan | Cross-sectional study | Two large Hospitals in the Chiba-prefecture, Japan (private and community hospitals)<br><br>2012-2016 | Claims data | Patients treated at either hospital during 2012-2016 meeting the claims based-algorithm<br><br>N= 3,294 | n= 400<br>Analysed= 389<br><br>Incident RA= 134 | F= 72.8% | 64.4 ± 14.9 | Prevalent and Incident RA | ICD-10 codes for RA (M05, M06)<br><br>Codes for relevant therapies and treatments | Medical records:<br>1. RA diagnosis by a physician<br>2. ACR/EULAR 2010 criteria<br>3. Overall adjudicator decision-confirmed cases |
| Thomas et al.<br>2008,<br>UK | Retrospective Cohort study | General Practice Research Database (GPRD), Primary care Medical Records, UK<br><br>1987-2002 | Electronic Primary Care Medical Records | Patients with RA or JIA registered in the GPRD<br><br>N=<br>RA= 31,830<br>JIA= 2,901 | RA=319<br>Analysed= 258<br><br>JIA= 101<br>Analysed= 91 | RA=<br>F= 71%<br><br>JIA=<br>F= 57% | RA=<br>Me: 51 (IQR 42-61)<br><br>JIA=<br>Me: 7 (IQR 4-10.5) | RA<br><br>JIA | RA and JIA codes<br>*(codes not specified)* | RA= 1987 ACR Criteria<br><br>JIA= 2001 ILAR Criteria<br><br>From AIS and Clinical Practices |

| | | | | | | | | | | |
|---|---|---|---|---|---|---|---|---|---|---|
| Waldenlind et al 2014,<br>Sweden | Retrospective Cohort study | National Patient Register and Prescribed Drug Register at the Karolinska University Hospital<br><br>2005-2012 | Hospital Discharge Records<br><br>Drug prescriptions | Patients with ≥1 primary or secondary ICD-10 code for seropositive or seronegative RA<br><br>N= NR | n= 211<br><br>Prevalent RA<br>n= 100<br>Analysed= 100<br><br>Incident RA<br>n=111<br>Analysed= 103 | Prevalent RA<br>F= 72%<br><br>Incident RA<br>F= 75% | Prevalent RA<br>57.9 ± 16.1<br><br>Incident RA<br>57.7 ± 17.4 | Prevalent and Incident RA | ICD-10 codes for RA (M05.9, M06.0) | Chart review from medical records<br><br>2010 ACR/EULAR criteria,<br>1987 ACR criteria or Clinical diagnosis |
| Wei et al. 2016,<br>USA | Cross-sectional study | Vanderbilt University Medical Center (VUMC), Tennessee<br><br>Administrative claims data<br><br>NR | Electronic Health Records<br>Clinical Notes<br><br>Billing codes | Patients identified in VUMC EHRs according to the 7 specified categories<br><br>N= NR | n= 175<br><br>(25 per each category) | NR | NR | RA | ICD-9<br><br>Clinical notes<br><br>Medication prescription | Manual chart review by 5 authors with medical background |
| Widdifield et al. 2013,<br>Canada | Retrospective Cohort study | 18 Ontario rheumatology clinics linked to health administrative data (Ontario Health Insurance Plan (OHIP) database, CIHI-DAD and NACRS)<br><br>1991-2011 | Physician billing codes<br><br>Hospital discharge records<br><br>Pharmacy data | ≥20 years active adults seen in rheumatology clinics with a visit between March 2009-2011 and seen at least twice<br><br>N= NR | n= 450<br>analysed= 447<br><br>RA=<br>n= 148<br><br>Non-RA=<br>n= 299 | RA=<br>F= 77.2%<br><br>Non-RA=<br>F= 64.8% | RA=<br>61.8 ± 14.1<br><br>Non-RA=<br>57.9 ± 64.8 | RA | ICD-9 and ICD-10 codes for RA (714, M05, M06)<br><br>Medication prescription | Medical records, clinical diagnosis by rheumatologists |
| Widdifield et al. 2014,<br>Canada | Retrospective Cohort study | 83 physicians in urban and rural Ontario linked to the Electronic Medical Record data Administrative Linked Database (EMRALD).<br><br>Ontario Health Insurance Plan (OHIP) Database, CIHI-DAD, NACRS and ODB<br><br>1991-2011 | Physician billing codes<br><br>Hospital discharge records<br><br>Pharmacy claims data | Patients seen in primary care clinics with at least 1 visit in the previous year (2010) and enrolled in the physician practice<br><br>N= 73,014 | ≥20 years:<br>n= 7,500<br><br>RA=<br>n= 69<br><br>non-RA=<br>n= 7431<br><br>≥65 years:<br>n= 3,426 | NR | ≥20 years:<br>49 ± 17 | RA | ICD-9 and ICD-10 codes for RA (714, M05, M06)<br><br>Medication prescription | Medical record review<br>1- diagnosis by rheumatologist, orthopaedic surgeon, or internal medicine specialist or,<br>2- primary care physician diagnosis with supporting evidence |

| | | | | | | | | | | |
|---|---|---|---|---|---|---|---|---|---|---|
| Zheng et al.[¥] 2022, USA | Case-control study | University of California, Los Angeles (UCLA) Health System<br><br>2009-2019 | Electronic Health Records (diagnosis, laboratory records) | RA:<br>≥18 patients from UCLA Health System with at least one ICD-9 or ICD-10 code<br><br>N= 22,266 and 2,385 confirmed with RA by clinician chart review<br><br>Non-RA:<br>Patients without ICD-9 or ICD-10 codes and with the exclusion criteria diagnosis (SLE and PA)<br><br>N= 2,381 | n=4,766<br><br>RA=2,385<br><br>Non-RA= 2,381 | RA= F= 83.1%<br><br>Non-RA= F= 70% | NR | RA | eMERGE algorithm<br><br>ICD-9 and ICD-10 codes for RA (714, 714.0, 714.1, 714.2, M05.*, M06.*<br><br>RF lab value<br><br>ICD-9 and ICD-10 codes for SLE and PA *(exclusion criteria)* | Clinician chart review<br><br>2010 ACR/EULAR criteria in a subsample (n=300) by rheumatologist expert, fellows and research associate |
| Zhou et al. 2016, UK | Retrospective Cohort study | CIPHER-SAIL databank (primary care general practice records linked with the rheumatology secondary care clinical system-Cellma) in ABMU and Cardiff regions<br><br>1999-2013 | Electronic Health Records (primary-Read codes- and secondary care-SNOMED-CT) | Patients >16 years registered in the general practice database contained in the SAIL databank and linked to Cellma system<br><br>*Developing set:*<br>ABMU cohort= N= 15,459<br><br>*Validation set:*<br>Cardiff cohort+ N= 5,208<br><br>N= 20,667 | n= 20,667<br><br>RA n= 2,588 | NR | NR | RA | Read codes and SNOMED-CT for RA, medications, and laboratory tests | Diagnosis of RA according to specialist rheumatologist recorded in Cellma |

Abbreviations: RA= Rheumatoid Arthritis; NR= Not Reported; NHS= National Health Service; CDs= Chronic Diseases; DMARDs= Disease-modifying Antirheumatic Drugs; JIA= Juvenile Idiopathic Arthritis; AIS= Additional Information Services; Me= Median; IQR= Interquartile Range; RF= Rheumatoid Factor; SLE= Systemic Lupus Erythematosus; PA= Psoriatic Arthritis; EMRs= Electronic Medical Records; BWH= Brigham and Women's Hospital; MGH= Massachusetts General Hospital; anti-CCP= antibodies to cyclic citrullinated peptide; VAMC= Veterans Affairs Medical Center;

THR= Total Hip Replacement; LiiRA= Lipids, Inflammation, and Cardiovascular risk in rheumatoid Arthritis; CPT= Current Procedural Terminology; CIHI-DAD= Canadian Institute for Health Information discharge Abstract Database; NACRS= National Ambulatory Care Reporting System; ODP= Ontario Drug Benefit Program; REP= Rochester Epidemiology Project; SADs= Systemic Autoimmune Diseases; CRANE= University of Nebraska Medical Center Clinical Research Analytics Environment; ILD= Interstitial Lung Disease; THA= Total Hip Arthroplasty; TKA= Total Knee Arthroplasty; AMI= Acute Myocardial Infarction; CIPHER-SAIL=Centre for Improvement in Population Health through E-Records- Secure Anonymised Information Linkage; ABMU= Abertawe Bro Morgannwg University.

^ Studies validating the same algorithm and same administrative data source.

¥ Studies validating the eMERGE algorithm

### Supplementary Appendix 5. Table 2

Table 2.1. Accuracy measures of individual studies assessing RA codes and algorithms in electronic health records (n=31).

| Author, Year, Country | Algorithm definition | Reference standard | Measures of Accuracy/Results<br>Sensitivity (95% CIs) | Specificity (95% CIs) | PPV (95% CIs) | NPV (95% CIs) | Other (95% CIs) |
|---|---|---|---|---|---|---|---|
| Allebeck et a. 1983, Sweden | Sole diagnosis RA (712.38 or 712.39 codes) | ≥3 New York criteria | NA | NA | 56.5%* | NA | |
| | | Diagnosis by rheumatologist | NA | NA | 65.2%* | NA | |
| Almutairi et al. 2021, Australia | ≥1 RA primary codes (M05.00–M06.99) | Rheumatologist diagnosis | 90% | 28.5% | 93.5% | 7.6% | |
| | | ACR/EULAR criteria | 89.8% | 16.6% | 80.5% | 30% | |
| | ≥1 RA primary biological infusion codes (I40Z) | Rheumatologist diagnosis | 25% | 71.4 | 90.9% | 7.7% | |
| | | ACR/EULAR criteria | 20.3% | 55.5% | 63.6% | 15.3% | |
| | ≥2 secondary RA secondary codes (M05.00–M06.99) | Rheumatologist diagnosis | 71.2% | 71.4% | 96.6% | 17.8% | |
| | | ACR/EULAR criteria | 71% | 44.4% | 83% | 28.5% | |
| | ≥2 RA primary codes (M05.00–M06.99) + biological infusion codes (I40Z) | Rheumatologist diagnosis | 60% | 85.7% | 97.9% | 15.8% | |
| | | ACR/EULAR criteria | 56.5% | 44.4% | 79.6% | 21% | |
| | ≥2 RA secondary codes (M05.00–M06.99) + biological infusion codes (I40Z) | Rheumatologist diagnosis | 76.2% | 57.1% | 95.3% | 17.4% | |
| | | ACR/EULAR criteria | 73.9% | 27.7% | 79.7% | 21.7% | |
| Carrara et al. 2015, Italy | *Step 1*: RA certification OR ICD 9-CM 714 in HDRs OR leflunomide OR tocilizumab OR abatacept OR gold salts | Medical Records clinically validated by an external investigator or according to specific classification criteria | 82.4% (77.6-86.5) | 96.2 % (94.2-97.7) | NR | NR | |
| | *Step 2*: Step 1 OR (methotrexate AND antimalarials AND no certification for other CADs) | | 85.4% (80.9-89.2) | 95.6% (93.5-97.2) | NR | NR | |
| | *Step 3*: Step 2 OR (GCs ≥3 prescriptions AND antimalarials AND no certification for other CADs | | 91.4% (87.6-94.3) | 92.2% (89.6-94.3) | NR | NR | |
| | *Step 4*: Step 3 OR (methotrexate ≥3 prescriptions AND no certification for other CADs (final model) | Training set | 96.3% (93.6-98.2) | 90.3% (87.4-92.7) | 85.0% (80.8-88.7) | 97.7% (95.9-98.9) | |
| | | Validation set-rheumatological sample | 93.7% (79.2-99.2) | 90.5% (81.5-96.1) | 81.1% (64.8-92.0) | 97.1% (89.9-99.6) | |
| | | Validation set- General population | 92.5% (79.6-98.4) | 99.8% (99.6-99.9) | 72.5% (58.3-84.1) | 99.9% (99.8-99.99) | |
| Carroll et al.¥ 2012, | *Algorithm (regression coefficient):* | Testing set- Partners Healthcare | 79% | NR | 88% | NR | ROC= 97% |

| | | | | | | | |
|---|---|---|---|---|---|---|---|
| USA | NLP RA (1.11) + NLP seropositive (0.74) + ICD-9 RA normalized 714.xx (0.71) + ICD RA 714.xx (0.66) + NLP erosions (0.46) + codified RF negative (0.36) + NLP methotrexate (0.3) + codified anti-TNF (0.29) + NLP anti CCP positive (0.27) + NLP anti-TNF (0.2) + NLP other DMARDs (0.13) – ICD JRA 714.3x (0.98) – ICD SLE 710 (0.57) – NLP PsA (0.51) | Testing set- EDW | 60% | NR | 87% | NR | ROC= 92% |
| | | Testing set- VUMC-SD | 57% | NR | 95% | NR | ROC= 95% |
| Cho et al.<br>2013<br>South Korea | RA code (M05.xx) + Biologics + Triple DMARDs + Any other DMARDs | Official report from a doctor documenting that the patient fulfils the 1987 ACR criteria | 96.5% | 61.9% | 93.3% | 76.2% | Accuracy: 91.2<br>PLR: 2.54 / NLR: 0.06 |
| | RA code (M05.xx) + Biologics + Triple DMARDs + Any NSAIDs | | 88.7% | 31.1% | 87.7% | 33.2% | Accuracy: 79.8<br>PLR: 1.29 / 0.36 |
| | RA code (M05.xx) + Biologics + Triple DMARDs + Any NSAIDs and any DMARDs | | 85.1% | 70.0% | 94.0% | 46.1% | Accuracy: 82.8<br>PLR: 2.84 / NLR: 0.21 |
| | RA code (M05.xx) + Biologics + Leflunomide + Any other DMARDs | | 96.5% | 61.4% | 93.2% | 76.2% | Accuracy: 91.1<br>PLR: 2.50 / NLR: 0.06 |
| | RA code (M05.xx) + Biologics + Leflunomide + Any NSAIDs | | 89.3% | 30.9% | 87.6% | 34.6% | Accuracy: 80.2<br>PLR: 1.29 / NLR: 0.35 |
| | RA code (M05.xx) + Biologics + Leflunomide + Any NSAIDs and any DMARDs | | 85.8% | 69.5% | 93.9% | 47.3% | Accuracy: 83.3<br>PLR: 2.81 / NLR: 0.20 |
| | RA code (M05.xx) + Biologics + MTX & Leflunomide + Any other DMARDs | | 96.5% | 60.6% | 92.9% | 76.2% | Accuracy: 90.8<br>PLR: 2.45 / NLR: 0.06 |
| | RA code (M05.xx) + Biologics + MTX & Leflunomide + Any NSAIDs | | 88.3% | 30.8% | 87.3% | 32.9% | Accuracy: 79.3<br>PLR: 1.28 / NLR: 0.38 |
| | RA code (M05.xx) + Biologics + MTX & Leflunomide + Any NSAIDs and any DMARDs | | 84.7% | 68.8% | 93.6% | 45.6% | Accuracy: 82.2<br>PLR: 2.72 / NLR: 0.22 |
| | Final: RA code (M05.xx) + Biologics + any DMARDs | | 96.5% | 58.7% | 92.3% | 76.2% | Accuracy: 90.3<br>PLR: 2.33 / NLR: 0.06 |
| Convertino et al.<br>2021,<br>Italy | RA according to HDRs or EDAs (ICD-9 code 714*) | Chart Medical Records; diagnosis by Rheumatologist | 0.53<br>(0.43-0.63) | 0.89<br>(0.83-0.93) | 0.74<br>(0.63-0.84) | 0.76<br>(0.70-0.82) | |
| | RA according to exemption code from co-payment (006) | | 0.77<br>(0.67-0.84) | 0.90<br>(0.85-0.94) | 0.82<br>(0.73-0.89) | 0.87<br>(0.81-0.91) | |
| | RA according to HDRs or EDAs (ICD-9 code 714*) AND RA according to exemption code from co-payment (006) | | 0.37<br>(0.28-0.47) | 0.95<br>(0.90-0.98) | 0.81<br>(0.67-0.91) | 0.72<br>(0.65-0.77) | |
| | RA according to HDRs or EDAs (ICD-9 code 714*) OR RA according to exemption code from co-payment (006) | | 0.93<br>(0.86-0.97) | 0.84<br>(0.78-0.90) | 0.78<br>(0.70-0.85) | 0.95<br>(0.91-0.98) | |
| Curtis et al.<br>2018<br>USA | ICD-9 codes 714.0, 714.2, 714.81 | Clinical diagnosis by rheumatologist in the Corrona registry | | | | | |
| | ≥1 year (Medicare) | | NR | NR | 0.70<br>(0.64-0.77) | NR | |
| | ≥1 year (Commercial Health Plan) | | NR | NR | 0.73<br>(0.58-0.89) | NR | |
| | ≥2 year (Medicare) | | NR | NR | 0.73<br>(0.64-0.82) | NR | |

| | | | | | | | |
|---|---|---|---|---|---|---|---|
| | ≥2 year (Commercial Health Plan) | | NR | NR | 0.68 (0.48-0.89) | NR | |
| | ICD-9 codes 714.xx | | | | | | |
| | ≥1 year (Medicare) | | NR | NR | 0.71 (0.64-0.77) | NR | |
| | ≥1 year (Commercial Health Plan) | | NR | NR | 0.74 (0.61-0.88) | NR | |
| | ≥2 year (Medicare) | | NR | NR | 0.75 (0.66-0.83) | NR | |
| | ≥2 year (Commercial Health Plan) | | NR | NR | 0.71 (0.52-0.91) | NR | |
| Fowles et al. 1995, USA | Any ICD-9-CM code for RA diagnosis in the claims | Medical record review | 44.4%* | 99.7%* | 44.4%* | 99.7%* | Agreement: 99.4% *K*= 0.44 |
| Hanly et al. 2015 Canada | A1: Two physician visits for RA at least 2 months apart | Medical chart review Rheumatologist's diagnosis | 83.0% (79.8-86.2) | 80.8% (79.1-82.5) | 51.9% (48.6-55.3) | 95.0% (94.0-96.0) | Accuracy: 81.2 (79.4-83.0) |
| | A2: A1, excluding individuals with at least 2 visits, at least 2 months apart, subsequent to the second RA visit, with 2 identical diagnosis of other IAs and CTDs and excluding diagnosis of RA when it was not confirmed by rheumatologist | | 68.8% (64.9-72.7) | 80.8% (79.1-82.5) | 47.2% (43.7-50.7) | 91.2% (90.0-92.5) | Accuracy: 78.3 (76.5-80.2) |
| | A3: 3 RA diagnostic billing codes over any time period | | 83.2% (80.0-86.3) | 81.6% (80.0-83.2) | 53.0% (49.7-56.4) | 95.1% (94.1-96.1) | Accuracy: 81.9 (80.2-83.7) |
| | A4: ≥1 hospitalisation where RA was in the diagnostic codes | | 20.7% (17.3-24.2) | 98.5% (97.9-99.0) | 77.1% (70.2-83.9) | 83.2% (83.2-84.7) | Accuracy: 82.9 (81.5-84.4) |
| | A5: ≥1 RA code by rheumatologist | | 87.7% (84.9-90.5) | 75.5% (73.7-77.3) | 47.2% (44.1-50.3) | 96.1% (95.1-97.0) | Accuracy: 77.9 (76.0-80.0) |
| | A6: A1 OR A5 OR A4 AND A2 | | 92.0% (89.7-94.3) | 74.3% (72.4-76.1) | 47.2% (44.1-50.2) | 97.4% (96.6-98.1) | Accuracy: 77.8 (75.8-79.8) |
| | A7: Any single diagnostic code for RA | | 94.8% (92.9-96.7) | 62.5% (60.4-64.5) | 38.7% (36.1-41.3) | 98.0% (97.2-98.7) | Accuracy: 68.9 (66.5-71.4) |
| Huang et al.¥ 2020 USA | *2010 Algorithm (regression coefficient):* NLP RA (0.970) + NLP seropositive (2.77) + ICD RA normalized 714.xx (66.0) + ICD RA 714.xx (0.639) + NLP erosions (1.26) + codified RF negative (0.851) + NLP methotrexate (0.632) + codified anti-TNF (0.959) + NLP anti CCP positive (1.31) + NLP anti-TNF (0.521) + NLP other DMARDs (0.298) – ICD JRA 714.3x (2.25) – ICD SLE 710 (0.959) – NLP PsA (0.856) | Validation set General RA mart (n=200) | 0.76 | 0.95 | 0.91 | 0.87 | ROC: 0.932 |
| | *2017 Updated Algorithm:* | Validation set General RA mart (n=200) | 0.77 | 0.95 | 0.91 | 0.87 | ROC: 0.937 |

| | | | | | | | |
|---|---|---|---|---|---|---|---|
| | Same model and regression coefficients incorporating ICD-10 codes (M05.x, M06.x) and updated medications to include newer anti-TNFs, anti-L6 and JAK inhibitors | Validation set<br>RA subjects with ICD-10 codes (no RA ICD-9 codes) (n=100) | 0.47 | 0.95 | 0.93 | 0.60 | ROC= 0.784 |
| | *Rule-based algorithm:*<br>≥3 ICD-9 RA codes | Validation set<br>General RA mart (n=200) | 0.79 | 0.59 | 0.54 | 0.82 | - |
| | *Rule-based algorithm:*<br>≥3 ICD-9 or ICD-10 RA codes | Validation set<br>General RA mart (n=200) | 0.89 | 0.58 | 0.56 | 0.90 | - |
| | ≥3 ICD-10 RA codes | Validation set<br>RA subjects with ICD-10 codes (no RA ICD-9 codes) (n=100) | 0.58 | 0.50 | 0.58 | 0.50 | - |
| Ibfelt et al.<br>2017<br>Demark | ICD-10 RA diagnostic codes (M05.9, M06.0, M06.8, M06.9) (DNPR= RA diagnosis in 2011 AND at least on additional visit with a RA diagnosis within 90 days after the first visit) | DANBIO (n= 573)<br>DNPR (n= 1,371) | NR<br>NR | NR<br>NR | 92%<br>79% | NR<br>NR | |
| Katz et al.<br>1997<br>USA | ICD-9-CM codes for RA (714.0, 714.1, 714.2, 714.3, 714.30, 714.31, 714.32, 714.33) | Medical records review-diagnosis by rheumatologists and 1990 ACR criteria | 0.90<br>(0.85-0.95) | NR | 0.95<br>(0.92-0.98) | NR | |
| Kim et al.<br>2011<br>USA | *No DMARD prescription filling required:*<br>≥2 claims associated with RA ICD-9-CM code 714 | RA by rheumatologist | NR | NR | 55.7%<br>(46.8-64.4) | NR | |
| | | ≥4 ACR criteria | NR | NR | 33.6%<br>(25.6-42.4) | NR | |
| | ≥3 claims associated with RA ICD-9-CM code 714 | RA by rheumatologist | NR | NR | 65.5%<br>(55.8-74.3) | NR | |
| | | ≥4 ACR criteria | NR | NR | 40%<br>(30.8-49.8) | NR | |
| | ≥2 RA claims by rheumatologist and separated by 7 days | RA by rheumatologist | NR | NR | 66.7%<br>(55.5-76.6) | NR | |
| | | ≥4 ACR criteria | NR | NR | 39.3%<br>(28.8-50.6) | NR | |
| | *At least 1 DMARD prescription filling is required*<br>≥2 claims associated with RA ICD-9-CM code 714 | RA by rheumatologist | NR | NR | 86.2%<br>(74.6-93.9) | NR | |
| | | ≥4 ACR criteria | NR | NR | 58.6%<br>(44.9-71.4) | NR | |
| | ≥3 claims associated with RA ICD-9-CM code 714 | RA by rheumatologist | NR | NR | 87.5%<br>(75.9-94.8) | NR | |
| | | ≥4 ACR criteria | NR | NR | 60.7%<br>(46.8-73.5) | NR | |
| | ≥2 RA claims by rheumatologist and separated by 7 days | RA by rheumatologist | NR | NR | 88.9%<br>(76.0-96.3) | NR | |
| | | ≥4 ACR criteria | NR | NR | 55.6%<br>(40.0-70.4) | NR | |

| | | | | | | | |
|---|---|---|---|---|---|---|---|
| Kronzer et al. 2020, USA | eMERGE RA algorithm: β = 1.937*log(1 + *RAICD*) + 1.639*_LabCount_ − 0.529*log(1 + *SLE*) − 0.122*log(1 + *Psoriatic*)− 0.954*log(1 + *Diagnoses*)^ | Manual chart review against 1987 ACR or 2010 ACR/EULAR criteria | 53% | 99% | 97% | 74% | ROC: 76% |
| | *RA duration:* 2 to <5 years | | 28% | 99% | 90% | 76% | ROC: 63% |
| | 5 to <10 years | | 63% | 97% | 94% | 80% | ROC: 80% |
| | ≥10 years | | 71% | 99% | 99% | 74% | ROC: 85% |
| | *Electronic Health Record lookback:* 1 year | | 13% | 100% | 100% | 61% | ROC: 56% |
| | 3 years | | 38% | 99% | 98% | 68% | ROC: 68% |
| | 5 years | | 45% | 99% | 96% | 71% | ROC: 72% |
| | 10 years and 15 years | | 52% | 99% | 97% | 73% | ROC: 75% |
| Kubota et al. 2021 Japan | 1A: RA code in one or more monthly claims | Chart review by rheumatologists | 86.3% (78-4-94.2) | 99.6% (99.5-99.7) | 56.8% (47.5-66.0) | 99.9% (99.9-100) | |
| | 1B: RA code in 1 inpatient or ≥2 outpatient monthly claims | | 83.6% (75.1-92.1) | 99.7% (99.6-99.8) | 58.7% (49.9-68.1) | 99.9% (99.9-100) | |
| | 2A: Any DMARD in ≥1 monthly claim | | 61.6% (50.5-72.8) | 99.9% (99.8-99.9) | 72.6% (61.5-83.7) | 99.8% (99.7-99.9) | |
| | 2B: Any DMARD in ≥2 monthly claims | | 60.3% (49.0-71.5) | 99.9% (99.8-99.9) | 74.6% (63.5-85.7) | 99.8% (99.7-99.9) | |
| | 3A: Oral corticosteroid in ≥1 monthly claim | | NA | NA | NA | NA | |
| | 3B: Oral corticosteroid in ≥2 monthly claim | | NA | NA | NA | NA | |
| | 4: SADs (other than RA) or Polymyalgia rheumatica (Exc.cri) | | NA | NA | NA | NA | |
| | 1A AND 2A | | 58.9% (47.6-70.2) | 99.9% (99.9-100) | 86% (76.4-95.6) | 99.8% (99.7-99.9) | |
| | 1A AND 2B | | 57.5% (46.2-68.9) | 100% (99.9-100) | 89.4% (80.5-98.2) | 99.8% (99.7-99.8) | |
| | 1B AND 2A | | 58.9% (47.6-70.2) | 100% (99.9-100) | 89.6% (80.9-98.2) | 99.8% (99.7-99.9) | |
| | 1B AND 2B | | 57.5% (46.2-68.9) | 100% (99.9-100) | 89.4% (80.5-98.2) | 99.8% (99.7-99.8) | |
| | 1A AND 3A | | 35.6% (24.6-46.6) | 99.8% (99.8-99.9) | 55.3% (41.1-69.5) | 99.6% (99.5-99.7) | |
| | 1A AND 3B | | 26% (16.0-36.1) | 99.9% (99.8-99.9) | 55.9% (39.2-72.6) | 99.6% (99.5-99.7) | |
| | 1A AND (2A OR 3A) | | 72.6% (62.4-82.8) | 99.8% (99.7-99.9) | 69.7% (59.4-80.1) | 99.8% (99.8-99.9) | |
| | 1A AND (2B OR 3B) | | 65.8% (54.9-76.6) | 99.9% (99.8-99.9) | 76.2% (65.7-86.7) | 99.8% (99.7-99.9) | |

| | | | | | | |
|---|---|---|---|---|---|---|
| | 1B AND (2A OR 3B) | | 67.1% (56.3-77.9) | 99.9% (99.8-99.9) | 77.8% (67.5-88.0) | 99.8% (99.7-99.9) |
| | 1A AND (2B or (3A AND 4)) | | 72.6% (62.4-82.8) | 99.9% (99.8-99.9) | 79.1% (69.4-88.8) | 99.8% (99.8-99.9) |
| | 1B AND (2B or (3A AND 4)) | | 72.6% (62.4-82.8) | 99.9% (99.8-100) | 80.3% (70.7-89.9) | 99.8% (99.8-99.9) |
| | 1B AND (2A or (3B AND 4)) | | 67.1% (56.3-77.9) | 99.9% (99.9-100) | 84.5% (75.2-93.8) | 99.8% (99.7-99.9) |
| | 2B OR (1B AND 3A AND 4) | | 75.3% (65.5-85.2) | 99.8% (99.7-99.9) | 70.5% (60.4-80.6) | 99.9% (99.8-99.9) |
| Liao et al. ¥ 2010 USA | *Algorithm (regression coefficient):* NLP RA (1.11) + NLP seropositive (0.74) + ICD-9 RA normalized 714.xx (0.71) + ICD RA 714.xx (0.66) + NLP erosions (0.46) + codified RF negative (0.36) + NLP methotrexate (0.3) + codified anti-TNF (0.29) + NLP anti CCP positive (0.27) + NLP anti-TNF (0.2) + NLP other DMARDs (0.13) – ICD JRA 714.3x (0.98) – ICD SLE 710 (0.57) – NLP PsA (0.51) | | | | | |
| | *Algorithm 1:* Narrative and codified (complete) | Validation set General RA Mart | 63% (51-75) | NR | 94% (91-96) | NR |
| | *Algorithm 2: Codified only* | | 51% (42-60) | NR | 88% (84-92) | NR |
| | *Algorithm 3: NLP only* | | 56% (46-66) | NR | 89% (86-93) | NR |
| Linauskas et al. 2018 Denmark | *Overall* Firs-time RA diagnosis registration ever in the DNPR (712.39, M05, M06) | Medical records, RA diagnosis verified against 1958 ACR, 1987 ACR or 2010 ACR/EULAR criteria. If unmet, clinical assessment by rheumatologist | NR | NR | 61.9% (56.6-67.0) | NR |
| | Firs-time RA diagnosis registration ever in the DNPR (712.39, M05, M06) AND ≥1 DMARDs prescription | | NR | NR | 87.7% (82.5-91.5) | NR |
| | *ICD-8 codes* Firs-time RA diagnosis registration ever in the DNPR | | NR | NR | 78.4% (61.5-89.2) | NR |
| | Firs-time RA diagnosis registration ever in the DNPR AND ≥1 DMARDs prescription | | NR | NR | 92.6% (72.8-98.3) | NR |
| | *ICD-10 M05 code* Firs-time RA diagnosis registration ever in the DNPR | | NR | NR | 62.0% (53.9-69.5) | NR |
| | Firs-time RA diagnosis registration ever in the DNPR AND ≥1 DMARDs prescription | | NR | NR | 80.2% (71.6-86.7) | NR |
| | *ICD-10 M06 code* Firs-time RA diagnosis registration ever in the DNPR | | NR | NR | 25.0% (18.5-32.8) | NR |
| | Firs-time RA diagnosis registration ever in the DNPR AND ≥1 DMARDs prescription | | NR | NR | 41.1% (30.2-52.9) | NR |

| | | | | | | | |
|---|---|---|---|---|---|---|---|
| Losina et al.<br>2003<br>USA | Codes 714 or 714.0 in Medicare (part A/hospital) claims | Medical record review by trained nurses | 0.65<br>(0.49-0.80) | NR | 0.86<br>(0.73-0.99) | NR | |
| | Codes 714 or 714.0 in Medicare (part A/hospital and part B/surgeon's) claims | | NR | NR | NR | NR | |
| Nanji et al.<br>2012<br>Canada | ICD-9 codes 714 or 714.0 | Chart review by rheumatologist clinical diagnosis | NA | NA | 70.9%* | NA | |
| Ng et al.<br>2012<br>USA | *All patients (with or without DMARDs therapy for ≥180 days*<br>2 RA codes (any visit) | Medical record review | NA | NA | 30.9%<br>(27.7-34.2) | NR | |
| | 2 RA codes + ≥1 RA code in rheumatologist visit | | 47.6<br>(41.4-53.8) | 68.3<br>(62.5-74.1) | 40.2<br>(34.1-46.3) | NR | |
| | 2 RA codes + ≥1 RA code in rheumatologist visit + last rheumatologist visit with and RA code | | 39.3<br>(30.5-48.0) | 91.5<br>(86.5-96.5) | 67.3<br>(58.9-75.8) | NR | |
| | *Patients with DMARD therapy for ≥180 days*<br>2 RA codes (any visit) | | 88.1<br>(84.7-91.5) | 74.1<br>(69.6-78.7) | 60.4<br>(55.3-65.5) | NR | |
| | 2 RA codes + ≥1 RA code in rheumatologist visit | | 44.6<br>(35.7-53.6) | 93.3<br>(88.9-97.8) | 75.0<br>(67.2-82.8) | NR | |
| | 2 RA codes + ≥1 RA code in rheumatologist visit + last rheumatologist visit with and RA code | | 38.1<br>(27.7-48.5) | 98.4<br>(95.7-100) | 91.4<br>(85.4-97.4) | NR | |
| Paltta et al.<br>2021,<br>Finland | ≥1 ICD 10 (M0.58, M05.9 or M06.0) code on CRHC visit with RA | All RA/chart review | NR | NR | 0.82<br>(0.76-0.87) | 1.00<br>(0.98-1.00) | PLR: 6.89 (5.21- 9.12)<br>NLR: 0.01 (0.00-0.04) |
| | | Sero(+) RA/chart review | NR | NR | 0.75<br>(0.67-0.83) | 1.00<br>(0.98-1.00) | PLR: 9.48 (6.71-13.4)<br>NLR: 0.01 (0.00-0.09) |
| | | Sero(-) RA/chart review | NR | NR | 0.71<br>(0.62-0.79) | 1.00<br>(0.98-1.00) | PLR: 8.44 (6.12-11.6)<br>NLR: 0.01 (0.00-0.10) |
| | ≥1 ICD 10 (M0.58, M05.9 or M06.0) code on CRHC visit with RA and reimbursement for DMARDs with inclusion diagnosis (202, 281, 313) | All RA/chart review | NR | NR | 0.89<br>(0.83-0.94) | 1.00<br>(0.98-1.00) | PLR: 16.4 (10.2-26.4)<br>NLR: 0.01 (0.00-0.05) |
| | | Sero(+) RA/chart review | NR | NR | 0.93<br>(0.84-0.98) | 1.00<br>(0.98-1.00) | PLR: 62.2 (23.5-164.4)<br>NLR: 0.02 (0.00-0.12) |
| | | Sero(-) RA/chart review | NR | NR | 0.79<br>(0.68-0.87) | 1.00<br>(0.98-1.00) | PLR: 16.3-10.1-26.2)<br>NLR: 0.02 (0.00-0.12) |
| | ≥1 ICD 10 (M0.58, M05.9 or M06.0) code on CRHC visit with RA and ACPA positivity | All RA/chart review | NR | NR | 0.98<br>(0.91-1.00) | 1.00<br>(0.98-1.00) | PLR: 246.03 (34.8-1740.2)<br>NLR: 0.02 (0.00-0.11) |
| | | Sero(+) RA/chart review | NR | NR | 0.96<br>(0.87-1.00) | 1.00<br>(0.98-1.00) | PLR: 123.2 (30.9-490.03)<br>NLR: (0.02 (0.00-0.13) |
| | | Sero(-) RA/chart review | NR | NR | - | - | - |
| Pedersen et al.<br>2004<br>Denmark | ICD-8 or ICD-10 codes for RA (712.19, 712.39, 712.59, M05, M06) | Clinical diagnosis (n=217) | NA | NA | 58.9% | NA | |
| | | 1987 ACR criteria (n=199) | NA | NA | 46% | NA | |

| | | | | | | | |
|---|---|---|---|---|---|---|---|
| Singh et al. 2004, USA | 1 ICD-10 code 714 alone | RA diagnosis by a rheumatologist on 2 separate occasions >6 weeks apart | 100% (NA) | 55% (48-62) | 66.2% (59-73) | 100% (NA) | ROC: 0.77 (0.69-0.85) |
| | ICD-10 714 and ≥3-month prescription of a DMARD | | 84.9% (80-90) | 82.7% (77-88) | 81.1% (75-87) | 86.2% (81-91) | ROC: 0.84 (0.77-0.90) |
| | ICD-10 code 714 and a positive RF | | 88.2% (83-93) | 91.4% (87-96) | 92.6% (88-97) | 86.5% (81-92) | ROC: 0.90 (0.84-0.95) |
| | ≥3-month prescription of a DMARD and positive RF | | 76.5% (70-83) | 95.7% (92-99) | 95.6% (92-99) | 77% (70-84) | ROC: 0.86 (0.80-0.92) |
| | ICD-10 code 714 and a DMARD prescription and positive RF | | 76.5% (70-83) | 97.1% (94-100) | 97% (94-100) | 77.3% (71-84) | ROC: 0.87 (0.81-0.93) |
| Sugiyama et al. 2024, Japan | *Prevalent group (n=389)*<br>One definite diagnosis within a claim month (M05, M06) AND no diagnosis of psoriasis (L40, L41, M07) AND prescription of any DMARD OR Glucocorticoid (n=389) | Physician diagnosis | NR | NR | 77.6% (73.5-81.8) | NR | |
| | | ACR/EULAR criteria | NR | NR | 22.6% (18.5-26.8) | NR | |
| | | Overall adjudicator decision-confirmed cases | NR | NR | 55.5% (50.6-60.5) | NR | |
| | One definite diagnosis within a claim month (M05, M06) AND no diagnosis of psoriasis (L40, L41, M07) AND any DAMRD (n=290) | Physician diagnosis | NR | NR | 88.6% (84.6-91.2) | NR | |
| | | ACR/EULAR criteria | NR | NR | 29.0% (23.7-34.2) | NR | |
| | | Overall adjudicator decision-confirmed cases | NR | NR | 71.7% (66.5-76.9) | NR | |
| | *Incident group (n=134)*<br>One definite diagnosis within a claim month (M05, M06) AND no diagnosis of psoriasis (L40, L41, M07) AND prescription of any DMARD OR Glucocorticoid (n=134) | Physician diagnosis | NR | NR | 59.7% (51.4-68.0) | NR | |
| | | ACR/EULAR criteria | NR | NR | 23.9% (16.7-31.3) | NR | |
| | | Overall adjudicator decision-confirmed cases | NR | NR | 38.8% (30.6-47.1) | NR | |
| | One definite diagnosis within a claim month (M05, M06) AND no diagnosis of psoriasis (L40, L41, M07) AND any DAMRD (n=78) | Physician diagnosis | NR | NR | 76.9% (67.6-86.3) | NR | |
| | | ACR/EULAR criteria | NR | NR | 41.0% (30.1-51.9) | NR | |
| | | Overall adjudicator decision-confirmed cases | NR | NR | 64.1% (53.5-74.8) | NR | |
| Thomas et al. 2008, UK | >1 RA code (on different dates) | 1987 ACR criteria applied from CPs records | 80% | 81% | 84%[*] | 76.2%[*] | OR: 16.8 (8.66-32.7)<br>AOR: 5.18 (2.06-12.9) |
| | RA diagnostic group 1 or 2[#] | | 93% | 49% | 69.9%[*] | 84.5%[*] | OR: 12.6 (5.77-27.7)<br>AOR: 6.33 (1.84-21.7) |
| | ≥1 DMARD prescription in GPRD with no prior alternative indication for the DMARD | | 78% | 96% | 96%[*] | 77%[*] | OR: 81.4 (27.5-241.02)<br>AOR: 55.5 (15.1-203.5) |

| | | | | | | | |
|---|---|---|---|---|---|---|---|
| | No alternative diagnosis on computer after last RA code | | 86% | 40% | 64.8%* | 69.5%* | OR: 4.20 (2.22-7.97)<br>AOR: 2.58 (0.94-7.11) |
| | An appropriate GPRD DMARD prescription OR no appropriate DMARD prescription and being in RA Group 1-2, having no alternative GPRD diagnosis for RA after last RA code and having >1 RA code during follow-up | | 84%<br>(73-94) | 86%<br>(72-92) | NR | NR | NR |
| Waldenlind et al<br>2014<br>Sweden | *Prevalent group (n=100)*<br>≥2 visits to rheumatologist between 2005-2008 listing an ICD-10 code M05.9 or M06.0 | 1987 ACR and 2010 ACR/EULAR criteria | NA | NA | 91%* | NA | |
| | *Incident group (n=102)*<br>First ever visit to a rheumatologist in 2008 listing an ICD-10 code M05.9 or M06.0 | | NA | NA | 83.3%* | NA | |
| Wei et al.<br>2016<br>USA | ICD-9 only | Manual chart review by 5 authors with medical background | 0.05 | NR | 0.36 | NR | F-score= 0.08 |
| | ≥2 ICD-9 codes | | 0.31 | NR | 0.77 | NR | F-score= 0.44 |
| | Primary Clinical notes only | | 0.60 | NR | 0.20 | NR | F-score= 0.30 |
| | Medications only | | 0.00 | NR | 0.00 | NR | F-score= 0.00 |
| | ICD-9 AND primary notes | | 0.25 | NR | 0.76 | NR | F-score= 0.38 |
| | ICD-9 AND medications | | 0.00 | NR | 0.64 | NR | F-score= 0.01 |
| | Primary notes AND medications | | 0.01 | NR | 0.88 | NR | F-score= 0.03 |
| | ICD-9 AND medications AND primary notes | | 0.08 | NR | 0.84 | NR | F-score= 0.15 |
| Widdifield et al.<br>2013,<br>Canada | *Patients ≥20 years (n=447)*<br>1 Hospitalisation (H) ever | Medical records, clinical diagnosis by rheumatologists | 23%<br>(16-30) | 96%<br>(94-99) | 76%<br>(63-88) | 72%<br>(67-76) | |
| | 1 H ever OR 1 Emergency Room (ER) visit ever | | 27%<br>(20-34) | 96%<br>(93-98) | 76%<br>(64-87) | 73%<br>(68-77) | |
| | 1 Physician diagnostic code (P) ever | | 100%<br>(100-100) | 60%<br>(54-65) | 55%<br>(49-61) | 100%<br>(100-100) | |
| | 1 P ever by a specialist | | 99%<br>(98-100) | 77%<br>(72-82) | 68%<br>(62-74) | 100%<br>(99-100) | |
| | (1 H ever) OR (2 P with ≥1 P by a specialist in 1 year) | | 98%<br>(96-100) | 82%<br>(78-86) | 73%<br>(67-79) | 99%<br>(97-100) | |
| | (1 H ever) OR (2 P with ≥1 P by a specialist in 2 years) | | 99%<br>(97-100) | 81%<br>(77-86) | 72%<br>(66-79) | 99%<br>(98-100) | |
| | (1 H ever) OR (2 P with ≥1 P by a specialist in 3 years) | | 99%<br>(97-100) | 81%<br>(76-85) | 72%<br>(65-78) | 99%<br>(98-100) | |
| | (1 H ever) OR (3 P with ≥1 P by a specialist in 1 year) | | 94%<br>(90-98) | 87%<br>(84-91) | 79%<br>(73-85) | 97%<br>(95-99) | |
| | (1 H ever) OR (3 P with ≥1 P by a specialist in 2 years) | | 97%<br>(94-100) | 85%<br>(81-89) | 76%<br>(70-82) | 98%<br>(96-100) | |
| | (1 H ever) OR (3 P with ≥1 P by a specialist in 3 years) | | 97%<br>(94-100) | 85%<br>(81-89) | 76%<br>(70-82) | 98%<br>(96-100) | |

| | | | | | | |
|---|---|---|---|---|---|---|
| | *Patients ≥65 years (n= 159)* | | 93% | 84% | 77% | 96% |
| | 1 P AND ≥1 DMARD or biologic agent | | (86-100) | (77-91) | (67-87) | (91-100) |
| | 2 P ≥60 days apart AND ≥1 DMARD or biologic agent | | 93% (86-100) | 88% (82-95) | 82% (72-91) | 96% (92-100) |
| | (1 H ever) OR (2 P AND ≥1 DMARD or biologic agent in 1 yr) | | 91% (84-99) | 92% (87-97) | 87% (78-95) | 95% (91-99) |
| | (1 H ever) OR (3 P AND ≥1 DMARD or biologic agent in 1 yr) | | 90% (82-97%) | 96% (92-100) | 93% (86-100) | 94% (90-99) |
| | (1 H ever) OR (3 P with ≥1 P by specialist AND ≥1 DMARD or biologic agent in 1 yr) | | 90% (82-97%) | 96% (92-100) | 93% (86-100) | 94% (90-99) |
| Widdifield et al. 2014, Canada | *Patients ≥20 years (n= 7500)* | Medical record review 1- diagnosis by rheumatologist, orthopaedic surgeon, or internal medicine specialist or, 2- primary care physician diagnosis with supporting evidence | 22% | 100% | 88% | 99% |
| | 1 Hospitalisation (H) ever | | (12-32) | (100-100) | (73-100) | (99-100) |
| | 1 H ever OR 1 Emergency Room (ER) visit ever | | 23% (13-33) | 100% (100-100) | 80% (63-98) | 99% (99-100) |
| | 1 physician diagnostic code (P) ever | | 90% (83-97) | 97% (96-97) | 20% (16-25) | 100% (100-100) |
| | 1 P ever by a specialist | | 81% (72-90) | 99% (99-100) | 51% (42-60) | 100% (100-100) |
| | 2 P by any physician in 1 year | | 84% (75-93) | 99% (99-99) | 46% (37-55) | 100% (100-100) |
| | 2 P by any physician in 2 years | | 84% (75-93) | 99% (99-99) | 45% (36-53) | 100% (100-100) |
| | 2 P by any physician in 3 years | | 84% (75-93) | 99% (99-99) | 42% (34-51) | 100% (100-100) |
| | 3 P by any physician in 1 year | | 80% (70-89) | 100% (99-100) | 63% (52-73) | 100% (100-100) |
| | 3 P by any physician in 2 years | | 80% (70-89) | 100% (99-100) | 60% (50-71) | 100% (100-100) |
| | 3 P by any physician in 3 years | | 80% (70-89) | 100% (99-100) | 59% (49-69) | 100% (100-100) |
| | 2 P with ≥1 P by specialist in 1 year | | 78% (69-88) | 100% (100-100) | 67% (56-77) | 100% (100-100) |
| | 2 P with ≥1 P by specialist in 2 years | | 78% (69-88) | 100% (100-100) | 67% (56-77) | 100% (100-100) |
| | 2 P with ≥1 P by specialist in 3 years | | 78% (69-88) | 100% (100-100) | 64% (54-75) | 100% (100-100) |
| | 3 P with ≥1 P by specialist in 1 year | | 77% (67-87) | 100% (100-100) | 82% (72-91) | 100% (100-100) |
| | 3 P with ≥1 P by specialist in 2 years | | 77% (67-87%) | 100% (100-100) | 80% (71-90) | 100% (100-100) |

| | | | | | | | |
|---|---|---|---|---|---|---|---|
| | 3 P with ≥1 P by specialist in 3 years | | 77%<br>(67-87%) | 100%<br>(100-100) | 80%<br>(71-90) | 100%<br>(100-100) | |
| | (1 H ever) OR (2 P with ≥1 P by specialist in 1 year) | | 80%<br>(70-89) | 100%<br>(100-100) | 66%<br>(56-76) | 100%<br>(100-100) | |
| | (1 H ever) OR (2 P with ≥1 P by specialist in 2 years) | | 80%<br>(70-89) | 100%<br>(100-100) | 66%<br>(56-76) | 100%<br>(100-100) | |
| | (1 H ever) OR (2 P with ≥1 P by specialist in 3 years) | | 80%<br>(70-89) | 100%<br>(99-100) | 64%<br>(54-74) | 100%<br>(100-100) | |
| | (1 H ever) OR (3 P with ≥1 P by specialist in 1 year) | | 78%<br>(69-88) | 100%<br>(100-100) | 79%<br>(79-89) | 100%<br>(100-100) | |
| | (1 H ever) OR (3 P with ≥1 P by specialist in 2 years) | | 78%<br>(69-88) | 100%<br>(100-100) | 78%<br>(69-88) | 100%<br>(100-100) | |
| | (1 H ever) OR (3 P with ≥1 P by specialist in 3 years) | | 78%<br>(69-88) | 100%<br>(100-100) | 78%<br>(69-88) | 100%<br>(100-100) | |
| | (1 H ever) OR (2 P ≥8 weeks apart in 2 years) | | 83%<br>(74-92) | 99%<br>(99-100) | 52%<br>(43-61) | 100%<br>(100-100) | |
| | *Patients ≥65 years (n= 3426)*<br>1 P AND 1≥ Rx (corticosteroids, DMARDs, biologics) ever | | 84%<br>(75-93) | 97%<br>(96-97) | 32%<br>(25-39) | 100%<br>(100-100) | |
| | 2 P AND 1≥ Rx (corticosteroids, DMARDs, biologics) ever | | 81%<br>(71-91) | 99%<br>(98-99) | 53%<br>(43-63) | 100%<br>(99-100) | |
| | (1 H ever) OR (2 P AND ≥1 Rx in 1 year) | | 83%<br>(73-92) | 99%<br>(99-100) | 67%<br>(56-77) | 100%<br>(100-100) | |
| | (1 H ever) OR (3 P AND ≥1 Rx in 1 year) | | 75%<br>(64-85) | 100%<br>(99-100) | 73%<br>(63-84) | 100%<br>(99-100) | |
| | (1 H ever) OR (3 P with ≥1 P by specialist AND ≥1 Rx in 1 year) | | 75%<br>(64-85) | 100%<br>(99-100) | 80%<br>(69-90) | 100%<br>(99-100) | |
| Zheng et al. 2022, USA | eMERGE RA algorithm:<br>$\beta = 1.937*\log(1 + \mathit{RAICD}) + 1.639*\mathit{LabCount} - 0.529*\log(1 + \mathit{SLE}) - 0.122*\log(1 + \mathit{Psoriatic}) - 0.954*\log(1 + \mathit{Diagnoses})^{\wedge}$ | Clinician chart review<br>2010 ACR/EULAR subset | 71.9%<br>74.5% | 95.2%<br>99.5% | 93.8%*<br>86.4%* | 77.2%*<br>74.4%* | F-score: 0.814 |
| Zhou et al. 2016, UK | *Phenotyping model (Decision tree) (8 code groups):*<br>- If number of occurrences of RARTH is 0 then return that RA is 0. If number of occurrences of RARTH is >0 then move to the next branch<br>- If INTENSITY_RA[#] is low (>2) then RA=0. If INTENSITY_RA is high (≤2) then move to the next branch of tree<br>- If evidence of PSORIATIC DIAGNOSIS >0 AND PREDNISOLONE prescription codes occurs ≤43 times then return RA=0, but if prednisolone codes occur >43 times then return RA | *Validation set*<br>Secondary care/GP overlap population in Cardiff (Cardiff-Cellma)<br>*Validation set*<br>Primary care population (ABMU-Cellma)<br>*Worst-case scenario (no record in outpatients signifies not RA)* | 86.2%<br><br><br><br>83% | 94.6%<br><br><br><br>99% | 85.6%<br><br><br><br>30.9% | NR<br><br><br><br>NR | Accuracy: 92.3% |

| | | | | | | |
|---|---|---|---|---|---|---|
| | - If no evidence of Psoriatic diagnosis AND evidence of METHOTREXATE OR SULPHASALAZINE (>18 prescriptions) OR LEFLUNOMIDE the return RA<br>- If no evidence of DMARDs but evidence of ALTERNATIVE_RA (alternative diagnosis) then return RA=0, if no evidence of alternative diagnosis then return RA=1 | *Best-case scenario (no record in outpatients signifies patient with RA treated elsewhere)* | 94% | 99% | 91.3% | NR |

Abbreviations: NR: Not Reported; NA= Not Applicable; CADs= Chronic Autoimmune Diseases; HDRs= Hospital Discharge Records; EDAs= Emergency Department Admissions; GCs= Glucocorticoids; RA= Rheumatoid Arthritis; CRCH= Finnish Care Register for Health Care; PLR: Positive Likelihood Ratio; NLR: Negative Likelihood Ratio; ACPA= Anti-citrullinated Protein Antibody; CPs= Clinical Practices; OR= Odds Ratio; AOR= Adjusted Odds Ratio; GPRD= General Practice Research Database; DMARDs= Disease-modifying Antirheumatic Drugs; RF= Rheumatoid Factor; ROC= Receiving Operating Characteristic curve area; NLP= Natural Language Processing; PsA= Psoriatic Arthritis; MTX: Methotrexate; EDW= Enterprise Data Warehouse; VUMC-SD= Vanderbilt University Medical Center's Synthetic Derivative; IAs= Inflammatory Arthritis; CTDs= Connective Tissue Diseases; DNPR= Danish National Patient Registry; RARTH= RA codes as defined in the NHS Wales QOF Rheumatoid Arthritis Indicator Set

[*] Calculated from data provided by 2x2 tables or when information was available.

[¥] Studies validating the same algorithm and same administrative data source.

[#] Four level of strength of evidence based on GPRD codes for potential RA cases, where 1= strongest evidence and 4= weakest evidence (Group 1 included codes for seropositive or erosive RA, Group 2 comprised "rheumatoid arthritis codes- such as RA of knee, Group 3 codes for systemic manifestations of RA and Group 4 codes for seronegative RA or other weak evidence for RA).

[^] *RAICD*, the total number of RA ICD-9 and ICD-10 diagnoses (714, 714.0, 714.1, 714.2, M05.*, M06. *); *LabCount*, whether the highest RF lab value seen in the patient's charts is either abnormal (LabCount = 1) or normal/unavailable (Lab Count = 0); *SLE*, the total number of SLE ICD-9/10 diagnoses; *Psoriatic*, the total number of psoriatic arthritis ICD-9/10 diagnoses; and *Diagnoses*, the total number of ICD-9/10 diagnoses in the patient's charts. Patients who scored greater than 0.632 were predicted as positive for RA.

Table 2.2. Accuracy measures of individual studies assessing JIA codes and algorithms in electronic health records (n=5).

| Author, Year, Country | Algorithm definition | Reference standard | Measures of Accuracy/Results<br>Sensitivity (95% CIs) | Specificity (95% CIs) | PPV (95% CIs) | NPV (95% CIs) | Other (95% CIs) |
|---|---|---|---|---|---|---|---|
| Harrold et al. 2013, USA | ≥1 diagnosis code from any provider | Manual chart review (Clinical diagnosis by adult or paediatric rheumatologist) | 100% | NR | 69% (59-78) | NR | |
| | ≥2 diagnosis codes from any provider | | 87% (76-93) | NR | 91% (80-96) | NR | |
| | ≥1 diagnosis code from rheumatology | | 81% (69-89) | NR | 90% (79-96) | NR | |
| | *Incident JIA*<br>≥1 diagnosis code from any provider | | 100% | NR | 45% (35-56) | NR | |
| | ≥1 diagnosis code from rheumatology | | 95% (84-99) | NR | 70% (57-81) | NR | |
| | ≥1 diagnosis code from rheumatology + ≥2 laboratory test performed (ANA, RF, HLA-B27) | | 93% (79-98) | NR | 75% (61-86) | NR | |
| | ≥1 diagnosis code from rheumatology + ≥2 laboratory test performed (ANA, RF, HLA-B27) + ≥12 months of pre-diagnostic enrolment | | 95% (81-99) | NR | 82% (67-91) | NR | |
| Peterson et al. 2023, USA | ≥1 ICD-9 codes alone | Chart review by paediatric rheumatologist (JIA ICD code at age ≤20 years and a rheumatology clinic note documenting diagnosis) | 82% | NR | 48% | NR | F-measure= 61% |
| | ≥2 ICD-9 codes alone | | 75% | NR | 68% | NR | F-measure: 71% |
| | ≥3 ICD-9 codes alone | | 72% | NR | 79% | NR | F-measure: 75% |
| | ≥4 ICD-9 codes alone | | 68% | NR | 82% | NR | F-measure: 75% |
| | ≥1 ICD-10 codes alone | | 76% | NR | 63% | NR | F-measure: 61% |
| | ≥2 ICD-10 codes alone | | 69% | NR | 76% | NR | F-measure: 73% |
| | ≥3 ICD-10 codes alone | | 72% | NR | 85% | NR | F-measure: 75% |
| | ≥4 ICD-10 codes alone | | 63% | NR | 88% | NR | F-measure: 74% |
| | ≥1 ICD-9 OR ICD-10 codes | Training set | 100% | NR | 48% | NR | F-measure: 64% |
| | ≥2 ICD-9 OR ICD-10 codes | | 93% | NR | 67% | NR | F-measure: 78% |
| | ≥3 ICD-9 OR ICD-10 codes | | 89% | NR | 78% | NR | F-measure: 83% |
| | ≥4 ICD-9 OR ICD-10 codes | | 87% | NR | 83% | NR | F-measure: 85% |
| | ≥1 ICD codes + “JIA” OR “JRA” | | 94% | NR | 67% | NR | F-measure: 78% |
| | ≥2 ICD codes + “JIA” OR “JRA” | | 88% | NR | 75% | NR | F-measure: 81% |
| | ≥3 ICD codes + “JIA” OR “JRA” | | 87% | NR | 84% | NR | F-measure: 86% |
| | ≥4 ICD codes + “JIA” OR “JRA” | | 86% | NR | 86% | NR | F-measure: 86% |
| | ≥1 ICD codes + “enthesitis” OR “uveitis” | | 80% | NR | 83% | NR | F-measure: 81% |
| | ≥2 ICD codes + “enthesitis” OR “uveitis” | | 79% | NR | 88% | NR | F-measure: 83% |
| | ≥3 ICD codes + “enthesitis” OR “uveitis” | | 77% | NR | 92% | NR | F-measure: 84% |

| | | | | | | | |
|---|---|---|---|---|---|---|---|
| | ≥4 ICD codes + "enthesitis" OR "uveitis" | | 78% | NR | 96% | NR | F-measure: 86% |
| | ≥1 ICD codes + "JIA" OR "JRA" AND "enthesitis" OR "uveitis" | | 78% | NR | 88% | NR | F-measure: 83% |
| | ≥2 ICD codes + "JIA" OR "JRA" AND "enthesitis" OR "uveitis" | | 77% | NR | 91% | NR | F-measure: 83% |
| | ≥3 ICD codes + "JIA" OR "JRA" AND "enthesitis" OR "uveitis" | | 77% | NR | 94% | NR | F-measure: 83% |
| | ≥4 ICD codes + "JIA" OR "JRA" AND "enthesitis" OR "uveitis" | | 77% | NR | 96% | NR | F-measure: 85% |
| | ≥4 ICD codes + "enthesitis" OR "uveitis" with exclusions (CTDs=710.0, 710.3, 710.4, M32, M33) | Training set | NR | NR | 97% | NR | F-measure: 87% |
| | | Validation set | NR | NR | 92% | NR | F-measure: 75% |
| | ≥4 ICD codes + "enthesitis" OR "uveitis" AND "JIA" OR "JRA" with exclusions (CTDs=710.0, 710.3, 710.4, M32, M33) | Training set | NR | NR | 97% | NR | F-measure: 86% |
| | | Validation set | NR | NR | 94% | NR | F-measure: 75% |
| | ≥4 ICD codes + "uveitis" with exclusions (CTDs=710.0, 710.3, 710.4, M32, M33) | Training set | NR | NR | 97% | NR | F-measure: 85% |
| | | Validation set | NR | NR | 92% | NR | F-measure: 75% |
| Shiff et al. 2017, Canada | ≥1 H or ≥2 diagnosis by any provider ever ≥1 day apart | Diagnosis by a paediatric rheumatologist | 90.8% (88.7-92.9) | 84.8% (81.3-88.3) | 91.1% (89.0-93.2) | NR | |
| | ≥1 H or ≥2 diagnosis by any provider ever ≥8 weeks apart | | 86.6% (84.0-89.0) | 88.7% (85.7-91.7) | 92.9% (90.9-94.9) | NR | |
| | ≥1 H or ≥2 diagnosis in 2 years by any provider ≥8 weeks apart | | 88.2% (85.7-90.7) | 90.4% (87.5-93.3) | 93.9% (92.0-95.8) | NR | |
| | ≥1 H or ≥2 diagnosis in 3 years by any provider ≥8 weeks apart | | 88.1% (85.6-90.6) | 89.7% (86.6-92.8) | 93.5% (91.5-95.5) | NR | |
| | ≥1 H or ≥2 diagnosis in 5 years by any provider ≥8 weeks apart | | 88.0% (85.4-90.6) | 88.2% (84.7-91.7) | 93.0% (90.9-95.1) | NR | |
| | ≥1 H or ≥3 diagnosis in 2 years by any provider ≥8 weeks apart | | 82.1% (79.2-85.0) | 92.6% (90.0-95.2) | 94.9% (93.1-96.7) | NR | |
| | ≥1 H or ≥3 diagnosis in 3 years by any provider ≥8 weeks apart | | 81.9% (78.9-84.9) | 92.1% (89.4-94.8) | 94.5% (92.6-96.4) | NR | |
| | ≥1 H or ≥3 diagnosis in 5 years by any provider ≥8 weeks apart | | 82.2% (79.1-85.3) | 91.8% (88.8-94.8) | 94.7% (92.8-96.6) | NR | |
| Stringer et al. 2015, Canada | 2 JIA codes (714) ≥8 weeks apart within 2 years by any physician | Clinical chart review (diagnosis by rheumatologist based on 2004 ILAR criteria) | 85% | NR | 60% | NR | |
| | 1 JIA code (714) by paediatric rheumatologist | | 86% | NR | 58% | NR | |
| | 2 JIA codes (714) ≥8 weeks apart within 2 years by paediatric rheumatologist | | 81% | NR | 65% | NR | |
| | 2 JIA codes (714) ≥8 weeks apart within 2 years by any physician excluding paediatric rheumatologist | | 53% | NR | 52% | NR | |
| Thomas et al. 2008, UK | >1 specific JIA code (on different dates) | 2001 ILAR criteria applied from CPs records (n=29) | 72% | 100% | 100%* | 36.4%* | |
| | >1 specific JIA code and/or nonspecific arthritis code | | 84% | 100% | 100%* | 50%* | |
| | 2 NSAID prescriptions in GPRD within 6 months of each other | | 96% | 100% | 100%* | 80%* | |

| | | | | | |
|---|---|---|---|---|---|
| | ≥1 DMARD prescription in GPRD record with no prior alternative indication for the DMARD | 40% | 100% | 100%* | 21%* |
| | No alternative diagnosis in GPRD record after last JIA code | 100% | 0% | 86.2%* | - |

Abbreviations: JIA= Juvenile Idiopathic Arthritis; JRA= Juvenile Rheumatoid Arthritis; ANA= Antinuclear Antibody; RF= Rheumatoid Factor; CTDs= Connective Tissue Diseases; H= Diagnosis in Hospital Discharge Records; CPs= Clinical Practices; NSAID= Non-steroidal Anti-Inflammatory Drugs; GPRD= General Practice Research Database; DMARDs= Disease-modifying Antirheumatic Drugs;

* Calculated from data provided by 2x2 tables when available.

### Supplementary Appendix 6. Figure 1

Figure 1. Forest Plot: individual and pooled estimates of sensitivity and specificity across algorithms categories (reference standard: diagnosis by rheumatologist).

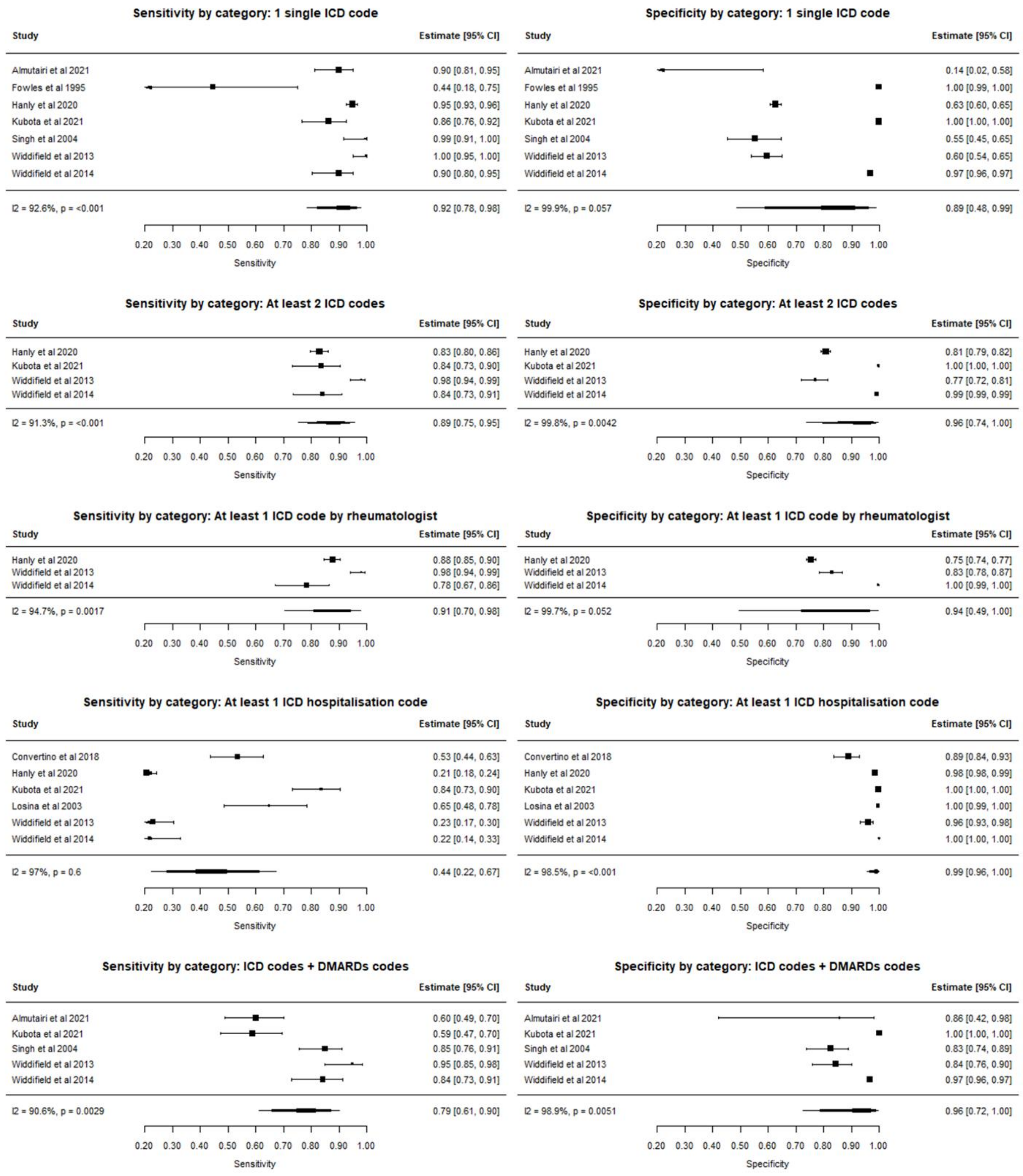

### Supplementary Appendix 7. Figure 2

Figure 2. Forest Plot: individual and pooled estimates of PPV across algorithms categories by reference standard (left: diagnosis by rheumatologist – right: ACR/EULAR classification criteria).

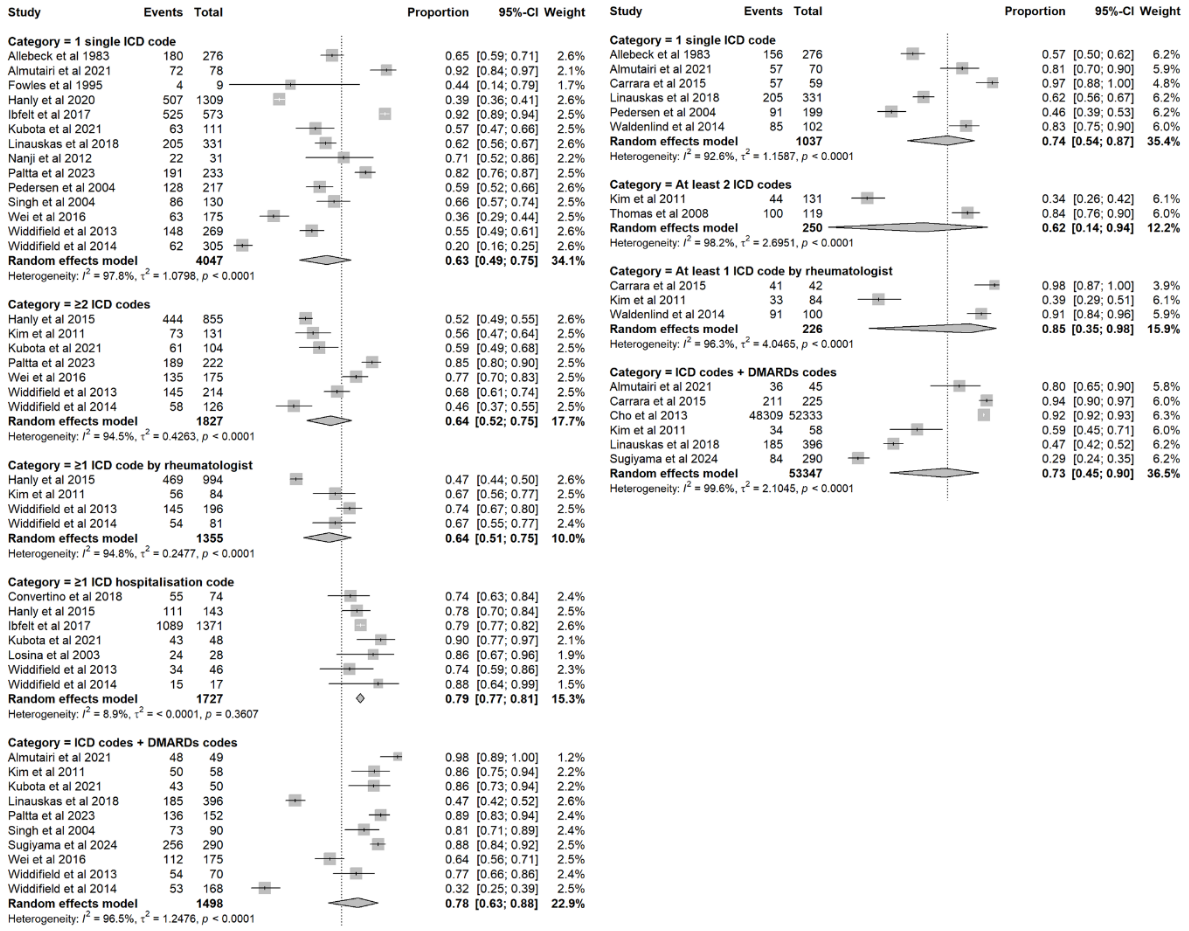
